# Systemic blockade of Clever-1 elicits lymphocyte activation alongside checkpoint molecule downregulation in patients with solid tumors

**DOI:** 10.1101/2020.11.11.20227777

**Authors:** Reetta Virtakoivu, Jenna Rannikko, Miro Viitala, Felix Vaura, Akira Takeda, Tapio Lönnberg, Jussi Koivunen, Panu Jaakkola, Annika Pasanen, Shishir Shetty, Maja de Jonge, Debbie Robbrecht, Yuk Ting Ma, Tanja Skyttä, Anna Minchom, Sirpa Jalkanen, Matti K. Karvonen, Jami Mandelin, Petri Bono, Maija Hollmén

## Abstract

Macrophages are critical in driving an immunosuppressive tumor microenvironment that counteracts the efficacy of T-cell targeting therapies. Thus, agents that can reprogram macrophages towards a proinflammatory state hold promise as novel immunotherapies for solid cancers. Here, we report that immunotherapeutic targeting of the macrophage scavenger receptor Clever-1 in heavily pretreated metastatic cancer patients was able to induce a significant increase and activation of peripheral T-cells. Anti-Clever-1 (FP-1305) administration led to suppression of nuclear lipid signaling pathways and a proinflammatory phenotypic switch in blood monocytes. Mechanistically, Clever-1 inhibition impaired multiprotein vacuolar ATPase–mediated endosomal acidification and improved macrophage cross-presentation of scavenged antigens. Our results reveal a non-redundant role played by the receptor Clever-1 in suppressing adaptive immune cell activation in humans. We provide evidence that targeting macrophage scavenging activity can promote an immune switch potentially leading to intratumoral proinflammatory responses in metastatic cancer patients.

## Introduction

Immunotherapies targeting CTLA-4 and PD-1/-L1 checkpoint molecules have revolutionized cancer treatment in the last few years and are now offering unforeseen prospects of cure for some patients. Despite these advances, the majority of patients remain refractory or develop resistance to these therapies for unspecified reasons (Sharma et al., 2017). In light of the current knowledge from prognostic and preclinical studies, tumor-associated macrophages are proving to be major contributors to therapeutic resistance by effectively suppressing antitumor immune responses (Cassetta and Pollard, 2018). Strategies under clinical development attempting to stimulate the antitumoral properties of these cells include Toll-like receptor (TLR), STING and CD40 agonists, inhibitors of PI3Kγ signaling and inhibitors of the CD47-SIRP*α* “don’t eat me” signaling axis that promotes phagocytosis of cancer cells (Jahchan et al., 2019). Preclinical studies have shown that in addition to these targets, Clever-1 (also known as Stabilin-1 and FEEL-1) shows potential in promoting macrophage reprogramming as it impairs the secretion of proinflammatory cytokines leading to ineffective activation of antigen-specific T-cell responses (Palani et al., 2016).

Clever-1 is a multifunctional scavenger and adhesion receptor expressed by human monocytes, subsets of immunosuppressive macrophages, lymphatic endothelial cells and sinusoidal endothelial cells in the liver and spleen. In macrophages, Clever-1 is involved in receptor-mediated endocytosis and recycling, intracellular sorting and transcytosis of altered and normal self-components (Hollmén et al., 2020). Immunotherapeutic targeting of Clever-1 in various tumor models delays tumor growth (Karikoski et al., 2014) by activating cytotoxic CD8^+^ T cells and renders refractory tumors more responsive to anti-PD-1 therapy (Viitala et al., 2019). To elucidate the significance of Clever-1 in suppressing antitumor immune responses in cancer patients, a phase I/II first-in-man clinical trial (MATINS; NCT03733990) was initiated to study the safety, tolerability and early efficacy of FP-1305, a humanized anti-Clever-1 antibody, in patients with selected advanced or metastatic solid tumors (Bono et al., 2020). FP-1305 antibody contains IgG4(S241P) heavy-chain and kappa light-chain constant regions and has further been optimized by introducing the L248E mutation to avoid Fcγ receptor binding (Reddy et al., 2000). Thus, FP-1305 has very low antibody-dependent cell cytotoxicity and complement-mediated effector functions, which is important to circumvent any unwanted effects on lymphatic and sinusoidal endothelial cells that express Clever-1 (Irjala et al., 2003).

In this study we describe the systemic immune signatures induced by FP-1305 administration in advanced cancer patients and provide a mechanistic understanding on how a macrophage-targeted approach can promote robust activation of T-cells and lead to promising antitumor immune responses.

## Results

### Clever-1 targeting promotes phenotypic conversion of circulating monocytes

Patients with various solid tumors (immunotherapy-refractory melanoma, pancreatic ductal adenocarcinoma, cholangio-, hepatocellular, ovarian and colorectal (CRC) carcinoma) received intravenous FP-1305 every three weeks in Part 1 of a phase I/II open-label trial. The tumor types were selected based on the association of Clever-1 expression (mRNA) with survival data obtained from the TCGA repository, analysis of Biobank material for macrophage Clever-1 positivity (Auria Biobank, University of Turku), and published research (Algars et al., 2012). Full clinical data were presented at ASCO 2020 virtual meeting (Bono et al., 2020). The drug was well-tolerated and no dose-limiting toxicity was observed during the dose-escalation phase with a maximum dose of 10 mg/kg.

In the circulation, Clever-1 is expressed by CD14^+^ classical monocytes (Palani et al., 2016), the precursors of tissue macrophages, and are the first to encounter intravenously administered FP-1305. In accordance, Clever-1 was most abundant on CD14^high^ monocytes in the study subjects and no positivity was observed on lymphocyte populations (Supplementary Fig. 1A). The cell surface Clever-1 expression on blood monocytes (CD14^high^) in the trial subjects (median fluorescence intensity 5965 range 13622) was not significantly different from healthy donors (4422 range 5741) (Supplementary Table 1). Cytometry time of flight (CyTOF) analysis of different myeloid populations at pre-dose (D0) and 7 days (D7) post FP-1305 dosing (**Fig. 1A, B**, Supplementary Fig. 1B and Supplementary Table 2) showed that FP-1305 induced a significant downregulation of CD206 and CD163 on CD14^high^ monocytes (**Fig. 1C**). CD206 and CD163 are the most common markers used to classify alternatively activated macrophages, which suggest that FP-1305 was able to reduce the immunosuppressive phenotype of circulating monocytes. Interestingly, the monocyte population increased in size and granularity by day 7 after treatment, during which we also observed a transient decrease in cell surface CD14 (**Fig. 1D**). FP-1305 did not induce depletion of Clever-1^+^ monocytes (detected by an alternate non-blocking antibody, 9-11) since the cell surface expression of Clever-1 remained constant or even increased during two weeks on the drug. However, the binding of FP-1305 to monocyte Clever-1 one day post-dose was nearly 100%, depicted as a drop in FP-1305-Alexa 647 detected Clever-1 signal. At day 7, the occupancy was cleared to some extent possibly because of the release of new monocytes from the bone marrow and trafficking of the targeted ones into tissues (**Fig. 1D**).

**Fig. 1.**
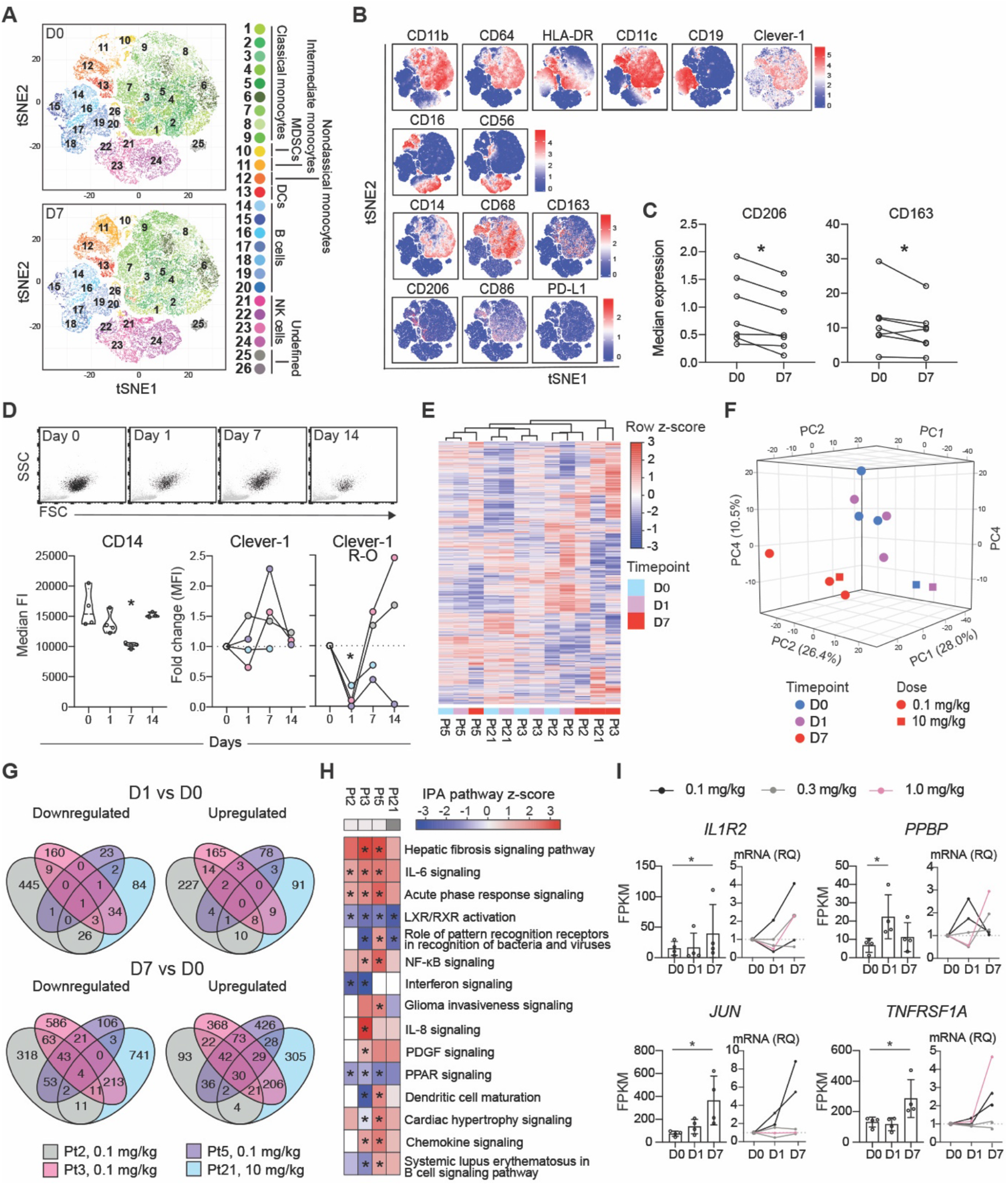
FP-1305 binds circulating CD14^+^ monocytes and suppresses LXR/RXR and PPAR signaling pathways. **A**, T-distributed Stochastic Neighbor Embedding (tSNE) of CD3-excluded circulating cells from pre-dose (D0, n=7) and day-7 (D7, n=7) samples pre-gated for viability, singlets and CD45^+^. **B**, tSNE heatmaps showing expression of indicated markers on cell clusters of a representative patient. **C**, Median expression of CD206 and CD163 on CD14^+^ monocytes at pre-dose and 7 days after FP-1305 treatment. Paired student’s *t*-test. **D**, Representative flow cytometry plots showing size (FSC) and granularity (SSC) of CD14^high^ monocytes (black), and median expression of CD14 and Clever-1 (9-11 ab) at different time points during treatment cycle 1. Clever-1 R-O signal was detected by alexa647 conjugated FP-1305 for investigating receptor (Clever-1) occupancy. One-way ANOVA performed between timepoints day 0, 1 and 7. **E**, Unsupervised hierarchical clustering of n=13,589 genes expressed in CD14^+^ monocytes obtained from four patients across timepoints day 0, 1 and 7. **F**, Principal component analysis of patient samples across different timepoints. **G**, Venn diagrams showing differentially expressed genes in comparison to pre-dose (D0). **H**, Ingenuity Pathway Analysis of patient gene expression changes at day 7 compared to pre-dose. Red color indicates predicted pathway activation and blue color inhibition. **I**, FPKM and RQ values of significantly altered genes at different timepoints. Friedman test. All *P* *<0.05.

### FP-1305 blocks tolerogenic gene expression related to LXR/RXR and PPAR pathways

To understand the treatment-induced monocyte activation we sequenced the CD14^+^ population 1 and 7 days after FP-1305 dosing. Unsupervised hierarchical clustering demonstrated a close relationship of the transcriptome between each patients’ pre and day 1 samples (**Fig. 1E**). However, the day 7 samples had more similarity by timepoint among the patients showing separation from other samples in principal component analysis (**Fig. 1F**) and overlapping gene expression changes (**Fig. 1G**). Ingenuity Pathway Analysis (IPA) of the differentially expressed genes between day 7 vs 0 revealed a significant downregulation of the LXR/RXR and PPAR pathways (**Fig. 1H**), possibly related to the FP-1305 induced impairment in modified LDL scavenging as shown previously in *ex vivo* cultured monocytes (Rantakari et al., 2016). At the same time, inflammation related pathways were activated as observed by a significant upregulation of *IL1R2, TNFRSF1A, JUN*, and *PPBP* (day 1) (**Fig. 1I**). These gene expression changes were verified by qPCR from the two sequenced patients (black) and expanded to other patient samples to show a rather heterogenous response related to dose and timepoint (**Fig. 1I**). It has to be emphasized that the several prior lines of therapies (Supplementary Table 1) might have influenced the release of new monocytes from the bone marrow, accounting for the observed differences in monocyte responses.

### Clever-1 harbors an interaction motif for the multiprotein vacuolar ATPase complex

To gain insight into FP-1305 uptake by Clever-1 on myeloid cells, dexamethasone polarized primary human macrophages were given conjugated forms of FP-1305(A647) and the non-competing antibody, 9-11(A488), for intracellular co-localization analysis. Both antibodies were rapidly internalized (Supplementary Fig. 2) and found to co-localize in intracellular vesicles. However, some structures were positive only for either one of the antibodies suggesting antibody-induced functional differences in endosomal sorting of Clever-1 (**Fig. 2A)**. To further explore this phenomenon, we performed coimmunoprecipitation (co-IP)/liquid chromatography-tandem mass spectrometry (LC-MS/MS) with FP-1305 and 9-11 (**Fig. 2B**). Co-IP/LC-MS/MS analysis of three biological replicates resulted in a list of 244 proteins enriched by either or both antibodies (Supplementary Data File). To identify and remove common background contaminants, we analyzed the list of 244 proteins against the CRAPome database with FC-A > 4 as the cut-off value for high-scoring interactions. This resulted in the removal of most nuclear, ribosomal and ribonucleoproteins and pruned the hits to 145 proteins of which 58 were specific for 9-11, 65 were specific for FP-1305 and 22 were common between the two antibodies (**Fig. 2C** and Supplementary Data File). We mapped all high-confidence (score > 0.7) protein–protein interactions reported in the STRING database between the 145 proteins in CytoScape and identified protein clusters with the MCL tool of the clusterMaker app (**Fig. 2D**). We used the STRING Enrichment tool to search for significant non-redundant (cut-off > 0.7) Gene Ontology Biological Process (GO BP) terms shared by proteins in the five largest clusters; protein transport and localization, RNA splicing, RNA translation, vesicle-mediated transport and phagocytosis (**Fig. 2D** and Supplementary Data File). We performed IPA separately on proteins precipitated with 9-11, proteins precipitated with FP-1305 and proteins that were immunoprecipitated with both antibodies. The most significant canonical pathway identified by IPA was phagosome maturation, which was obtained with 9-11 (**Fig. 2E**). The proteins constituting this pathway were subunits of the lysosomal membrane V0 and the cytosolic V1 sectors of the vacuolar ATPase (v-ATPase) (**Fig. 2F**). Specificities of the v-ATPase subunits ATP6V1A, ATP6V0A1 and TCIRG1 for the non-interfering 9-11 antibody were validated by co-IP/western blot (**Fig. 2G**). Since FP-1305 was unable to precipitate Clever-1 when bound to these subunits, it can be postulated that FP-1305 can interfere in the acidification of phago-lysosomes during phagosome maturation.

**Figure 2.**
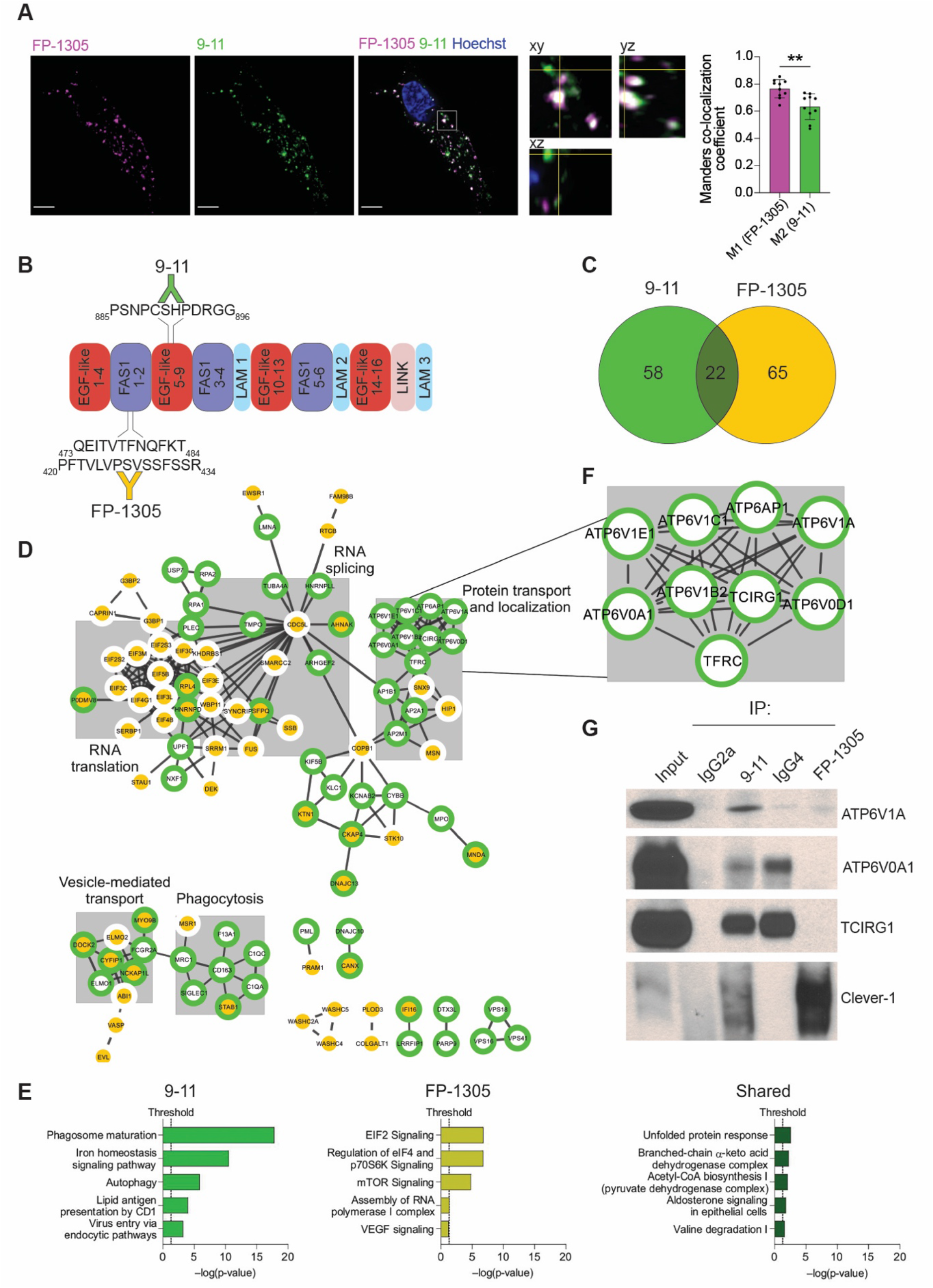
Clever-1 harbors an interaction motif for the multiprotein vacuolar ATPase complex. **A**, Representative confocal images of FP-1305 and 9-11 internalization into primary human macrophages after 2h antibody treatment (n = 2 healthy donors). A single focal plane and orthogonal view of the indicated magnified area show vesicles containing either one or both of the antibodies. Co-localization was assessed with Manders’ co-localization coefficients M1 (FP-1305) and M2 (9-11), n=10 cells from two independent experiments with the same donor. Scale bars 5μm. Statistical comparison of co-localization was conducted using Wilcoxon matched-pairs signed rank test. ** *p* <0.01. **B**, Schematic of the main 9-11 and FP-1305 antibody epitopes mapped on the human Clever-1 protein primary structure: EGF-like, epidermal-growth-factor-like domain; FAS1, fasciclin domain; LAM, laminin-type EGF-like domain; LINK, C-type lectin-like hyaluronan-binding LINK module. **C**, Venn diagram showing the number of 9-11-specific and FP-1305-specific proteins and proteins shared by both antibodies in the CRAPome-pruned Clever-1 interactome. **D**, High-confidence protein– protein interactions between proteins in the Clever-1 interactome retrieved from the STRING database and mapped in Cytoscape. Proteins present in the 9-11 interactome have a green border and proteins present in the FP-1305 interactome a yellow center. The five largest clusters are highlighted with grey boxes and titled after common themes in their significantly enriched GO Biological Process terms. **E**, Significantly enriched pathways from IPA performed separately on 9-11-specific, FP-1305-specific and shared proteins. **F**, Magnification of the Clever-1 interactome showing subunits of the v-ATPase complex, which immunoprecipitated specifically with 9-11. **G**, Co-immunoprecipitation/western blot validation showing that the v-ATPase subunits ATP6V1A, ATP6V0A1 and TCIRG1 are immunoprecipitated with 9-11 but not with FP-1305. Clever-1 signal was detected with 3-372 antibody, which is the parent antibody of FP-1305.

### Clever-1 is required for efficient lysosomal acidification and degradation of antigens

The functionally active v-ATPase is an ATP-dependent proton pump that regulates lysosomal acidification through the reversible assembly of its V0 and V1 sectors on the lysosomal surface (Collins and Forgac, 2020). To investigate the functional effects of the observed v-ATPase and Clever-1 interaction, we utilized the acute myelogenous leukemia cell line KG-1, which expresses high levels of Clever-1 and differentiates into macrophage-like cells after PMA treatment (Ackerman and Cresswell, 2003; St Louis et al., 1999; Teobald et al., 2008). We used RNA interference to knock down Clever-1 expression (**Fig. 3A**) and kinetically monitored steady-state total cellular acidification and DQ-OVA antigen degradation in KG-1 macrophages. Clever-1 knockdown significantly impaired cellular acidification and antigen degradation (**Fig. 3B, C**). To more specifically interrogate the contribution of Clever-1 to these processes, we performed endocytosis and acidification experiments with acetylated LDL (acLDL), a ligand of Clever-1 and other scavenger receptors. Predictably, Clever-1 knockdown significantly reduced but did not completely inhibit acLDL endocytosis (**Fig. 3D**). However, Clever-1 knockdown significantly impaired the acidification of endocytosed acLDL (**Fig. 3E**). These results were independently verified using another siRNA (Supplementary Fig. 3). Confocal microscopy further verified the co-localization of Clever-1 and ATP6V0A1 in LAMP-1^+^ lysosomes and demonstrated an increase in the quantity of lysosomal ATP6V0A1 after acLDL treatment (**Fig. 3F, G, H**). Treating primary human macrophages with FP-1305 led to an increase in cellular pH similar to Clever-1 knockdown (**Fig. 3I**). It has been suggested that reduced cellular acidification rescues antigen from degradation to cross-presentation (Alloatti et al., 2015). To analyze how Clever-1 deficiency affects antigen cross-presentation, we used bone-marrow-derived macrophages from wildtype and Clever-1 knockout (Clever-1^−/−^) mice that were TAM-polarized *in vitro* and fed ovalbumin (OVA) SIINFEKL peptide, full-length OVA protein or cell-associated OVA on apoptotic EG.7 cells. Clever-1^−/−^ macrophages cross-presented higher levels of antigen compared to wildtype macrophages after all treatments (**Fig. 3J**).

**Figure 3.**
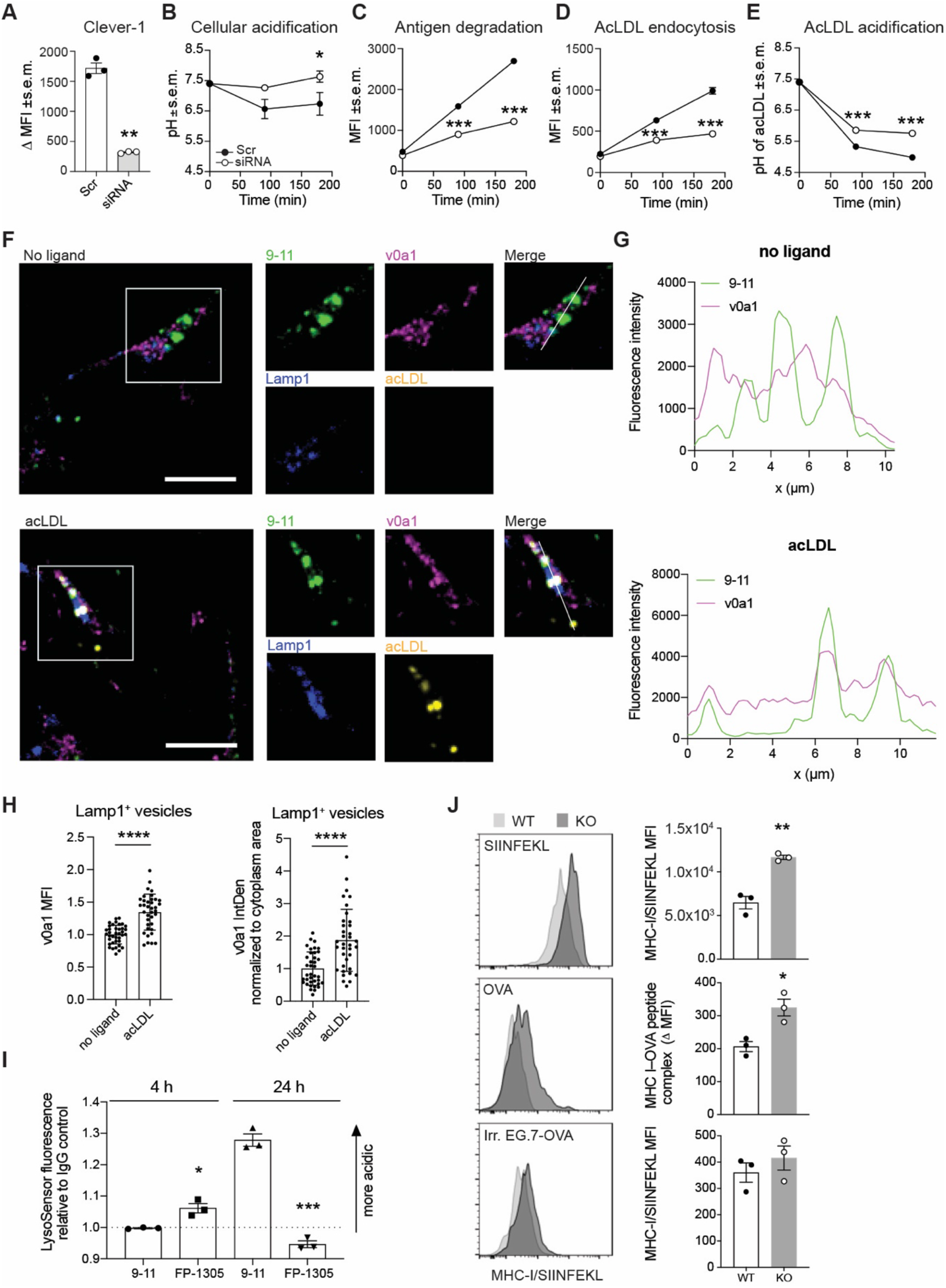
Clever-1 regulates the acidification and degradation of endocytosed antigens. **A**, Clever-1 knockdown efficiency in KG-1 macrophages quantified by flow cytometry with 9-11 antibody; n=3, Student’s paired two-tailed *t*-test. **B–E**. Total cellular acidification (**B**), antigen degradation (**C**), acLDL endocytosis (**D**) and acLDL acidification (**E**) kinetics in KG-1 macrophages transfected with scramble (Scr) or Clever-1 siRNA; n=3, Two-way ANOVA with Sidak’s multiple comparisons test (B-E). **F**, Confocal microscopy images of Clever-1 (green), ATP6V0A1 (magenta) and LAMP-1 (blue) localization in KG-1 macrophages incubated with or without 10 μg/ml acLDL (yellow) for three hours. Scale bars 10 μm. **G**, Intensity profiles generated from confocal images of Clever-1 (green) and ATP6V0A1 (magenta) in KG-1 macrophages incubated with or without 10 μg/ml acLDL (yellow) for three hours. Intensities obtained across the white lines in the corresponding merged images. **H**, ATP6V0A1 mean fluorescence intensity (MFI) and integrated density (IntDen) in LAMP1^+^ lysosomes of KG-1 macrophages treated with and without 10 μg/ml acLDL, n > 35 cells from three independent experiments normalized to each experiment’s no ligand -mean. **I**, M2-polarized primary human macrophages from three donors were treated with 50 ng/ml of 9-11 or FP-1305 for the indicated time points. Changes in cellular pH were measured by LysoSensor Green fluorescence intensity relative to cells treated with control IgG. Student’s un-paired two-tailed *t*-test. **J**, Flow cytometric detection of cross-presented ovalbumin (OVA) on LLC (Lewis lung carcinoma)-conditioned medium polarized peritoneal Clever-1 wild type (WT) or knockout (KO) macrophages after treatment with OVA peptide, OVA protein or apoptotic EG.7 cells. Student’s un-paired two-tailed *t*-test. * *p* <0.05, ** *p* < 0.01, *** *p* < 0.001. All error bars denote SEM.

### FP-1305 induces robust T-cell activation and downregulation of immune checkpoint expression

To investigate how the FP-1305-induced changes in monocyte and macrophage function was contributing to adaptive immune responses in vivo we analyzed lymphocyte populations in MATINS study subject pre- (D0) and day 7 (D7) post-treatment samples with CyTOF. Unbiased PhenoGraph clustering of lymphocytes identified 22 different T-cell (CD3^+^) metaclusters across all patient samples (**Fig. 4A, B** and Supplementary Table 3). The analysis showed changes in several T-cell populations across patients receiving different doses of FP-1305 (1, 3 or 10 mg/kg), indicating that this transition was not dose-dependent (**Fig. 4C**). The most pronounced effects were seen within the naïve CD4^+^ (#5), naïve (#11) and effector (EFF) CD8^+^ (#15) clusters with increased CXCR3 and CD25 expression, respectively (**Fig. 4C, D** and Supplementary Fig. 4). In addition, we observed a significant induction of a CD8^+^ effector memory (EM) population (#18) with upregulation of CD25 and CXCR3 (**Fig. 4C, D**). In contrast, CD25 was not significantly upregulated in the regulatory T-cell population (Treg, cluster #10) (**Fig. 4D**). In T-cells, CD25 (interleukin-2 receptor alpha chain) expression is induced by interleukin-2 few days after T-cell receptor (TCR) ligation enabling clonal expansion of antigen experienced cells. Consequently, CXCR3 expression is rapidly induced on naïve T-cells and facilitates their migration into the tumor tissue and differentiation into effector cells (De Simone et al., 2019). The upregulation of activation markers in most clusters occurred regardless of the changes in cluster size, proposing a general induction of peripheral T-cell activation (**Fig. 4D**). In support of this, the expression of several regulatory immune checkpoint molecules CTLA-4, LAG-3, PD-1 and PD-L1 were downregulated on CD4^+^ T-cell populations (**Fig. 4D**). The expression of the co-stimulatory receptor CD28 but not CD27 was also diminished on activated (naïve) CD4^+^ T-cells possibly suggesting a transition to terminally differentiated effector cells (Mou et al., 2014). The peripheral T-cell activation was accompanied by increased Ki67 expression in CD8^+^ EFF (in 5/6 of the patients), EM (4/6) and double-negative (CD4^-^CD8^-^) (5/6) T-cells indicative of their proliferation (**Fig. 4E**). Moreover, the double-negative and CD8^+^ T-cells together with NKT-cells produced increasing levels of perforin 7-14 days after FP-1305 dosing (**Fig. 4F**) providing evidence of their higher cytotoxic potential. In addition, we observed increased secretion of IL-2 and interferon (IFN)γ in peripheral CD8^+^ T-cells 7-14 days after treatment (**Fig. 4G**).

**Fig. 4.**
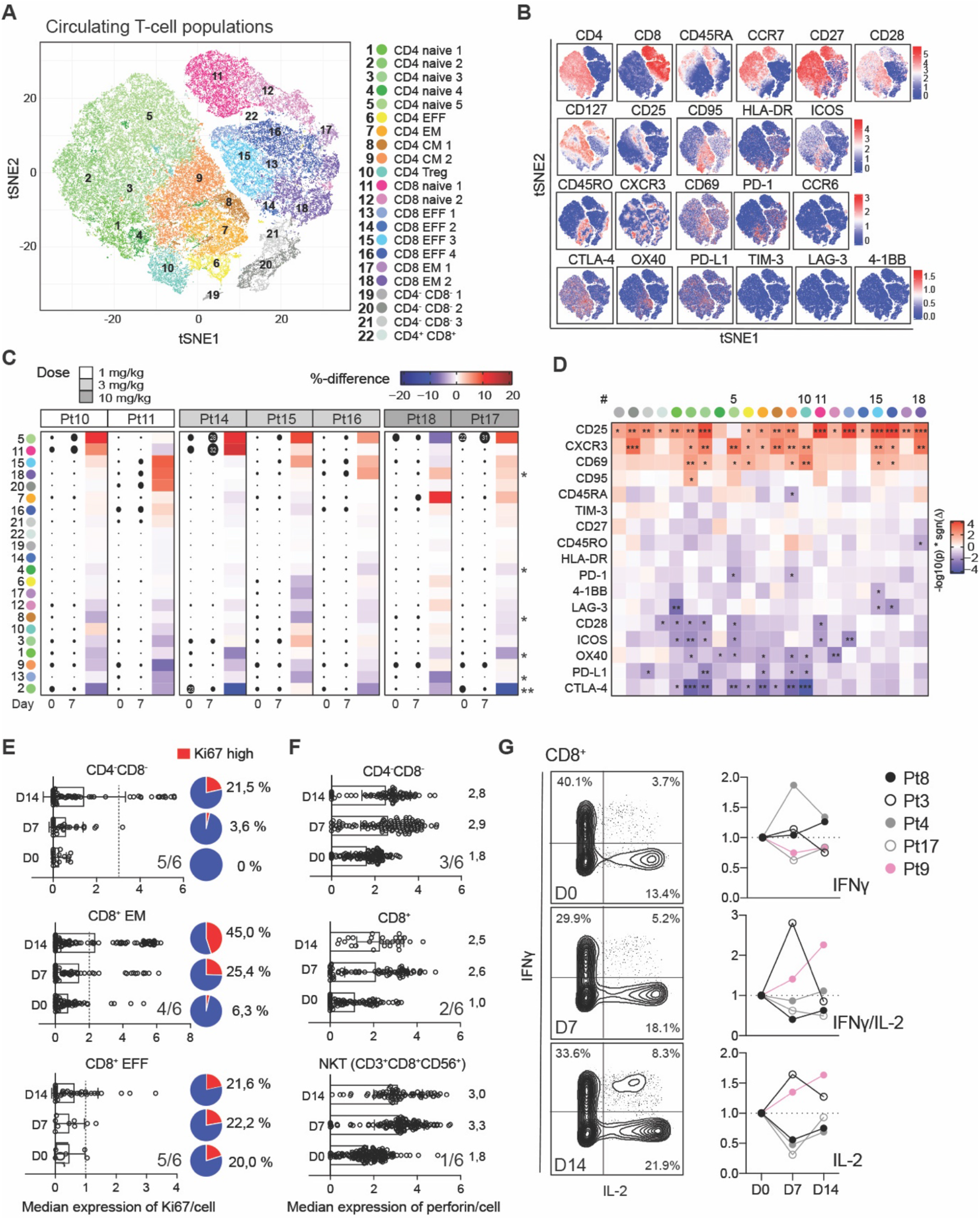
FP-1305 elicits activation and proliferation of circulating cytotoxic T-cells. **A**, tSNE plots of circulating T-cells pre-gated for viability, singlets and CD3^+^, 5000 cells/sample (n=7). EFF, effector; EM, effector memory; Treg, regulatory T-cell. **B**, tSNE heatmaps showing expression of indicated markers on T-cell clusters of a representative patient. **C**, Heatmap of cluster size pre- (D0) and post- (D7) treatment. Dot size indicates the relative percent of the cluster per sample. Red color points to positive and blue to negative change between pre- and post-samples within a patient. The asterisk indicates statistical significance of the change across all patients. **D**, Heatmap of marker differences between pre- and post-samples in each T-cell cluster (n=7), paired student’s *t*-test (C, D). **E**, Median expression of Ki67 and **F**, perforin in selected T-cells from a representative patient (Pt10). Overall response across six patients with at least >10% increase in Ki67 and perforin is indicated in the right corner of each graph. Cut-off for Ki67^high^ is depicted as a dashed line for each cell type. The median expression of perforin is indicated for each timepoint on the right side of the bars. **G**, Expression of IFNγ and IL-2 on PMA/ionomycin stimulated peripheral CD8^+^ T-cells obtained at pre-dose or 7 and 14 days after FP-1305 dosing. Plots are shown for patient Pt9 (pink). * *p* <0.05, ** *p* < 0.01, *** *p* < 0.001

### FP-1305 restores patient responses to pro-inflammatory stimulus

IFNγ is critical for T-, NKT- and NK-cell trafficking into tumors via induction of CXCR3 ligands CXCL9, CXCL10 and CXCL11 (Melero et al., 2014). Therefore, we measured the change of systemic IFNγ and CXCL10 in FP-1305 treated patients across different dosing cycles. Seven (23%) and six (20%) out of the 30 patients showed more than a two-fold increase in IFNγ and CXCL10 by two weeks on treatment, respectively (**Fig. 5A**). Such a response was seen at all dose levels, although most frequently at dose levels 0.3 and 1 mg/kg. Since most cancer patients have a Th2-skewed immune profile we wanted to test whether FP-1305 can reinvigorate patient responses to TLR4 mediated activation. Indeed, peripheral blood mononuclear cells (PBMCs) isolated from patients after having received FP-1305 for 24 hours showed improved IFNα and β secretion after *ex vivo* LPS exposure compared to their PBMCs from pre-dose samples (**Fig. 5B**). At the same time, the shedding of sCD163 was increased in post treatment samples supporting increased immune activation not only of T- and NK-cells but also of peripheral monocytes (Etzerodt et al., 2014) further supporting the CyTOF and RNA-seq data.

**Fig. 5.**
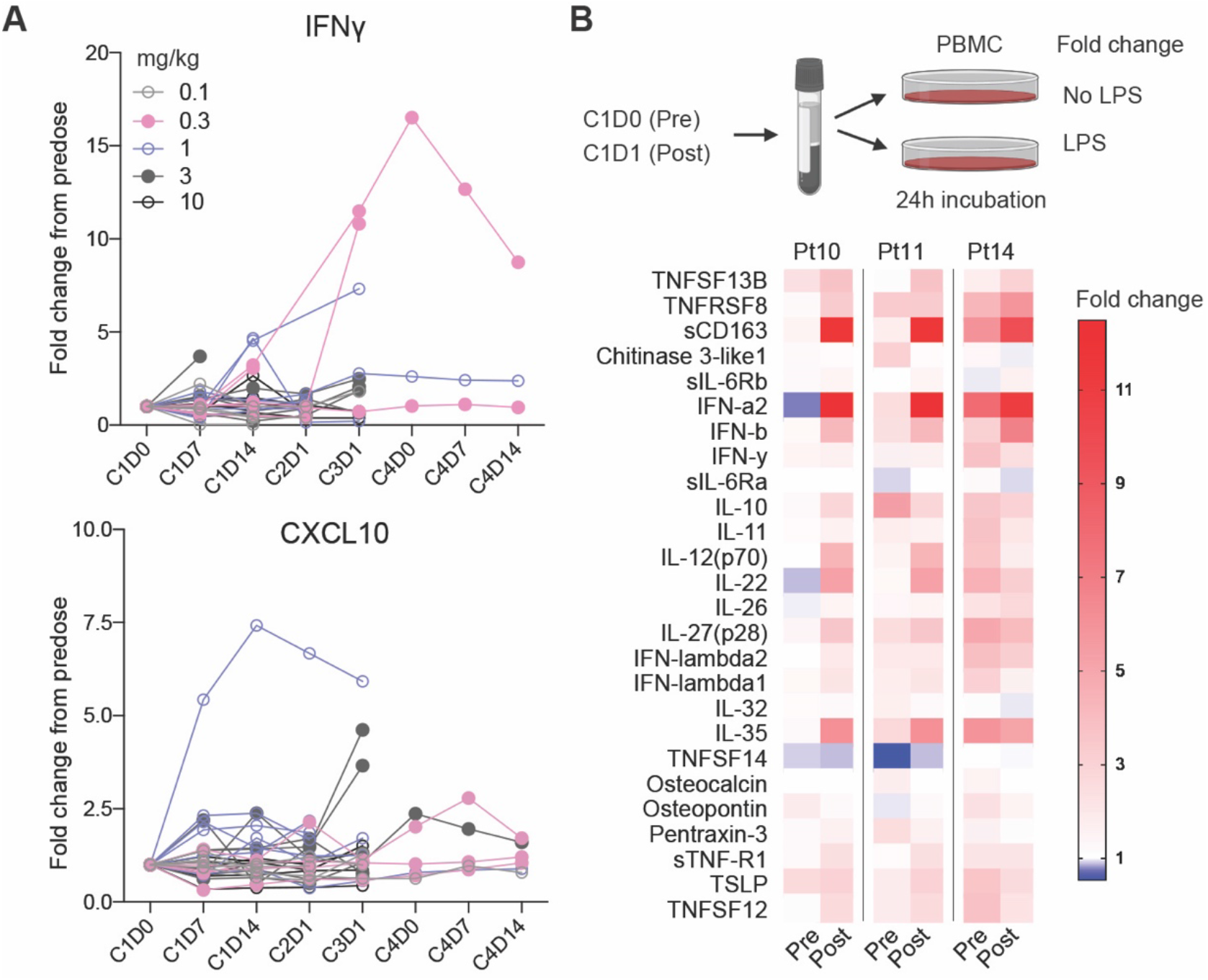
FP-1305 re-invigorates patient responses to inflammatory stimulus by increasing the secretion of interferons. **A**, Systemic IFNγ and CXCL10 in MATINS trial patients during dosing cycles (C) 1-4 shown as fold-change from baseline. **B**, Schematic of *ex vivo* LPS (TLR4 specific) stimulation of patient PBMCs 24 hours after receiving the first dose of FP-1305 and heatmap of LPS-induced secretion of different factors at pre- and post-dose shown as fold change from unstimulated cells. *P* *<0.05, **<0.01, ***<0.001.

### FP-1305 promotes clonal expansion of CD8 effector T-cells

To understand the relevant peripheral changes that would be indicative of an FP-1305 induced effective anti-tumor response we looked more closely on data obtained from a CRC patient with a partial response according to Response Evaluation Criteria in Solid Tumors Version 1.1 (RECIST 1.1) (Eisenhauer et al., 2009) (**Fig. 6A)**. Immunohistochemical staining of the primary tumor showed very high number of Clever-1 and CD163 positive macrophages and few intratumoral CD8^+^ T-cells (**Fig. 6B**). FP-1305 dosing induced a circulating CD8^+^ effector T-cell population expressing CD69, CD25, CXCR3, and ICOS, which was in the patient’s case most effectively induced at cycle 4 (**Fig. 6C**). We used the cycle 4 sample to identify the same T-cell population from the other patients previously analyzed with CyTOF, and found that already during cycle 1, this population increased substantially at day 7 (**Fig. 6D**).

**Fig. 6.**
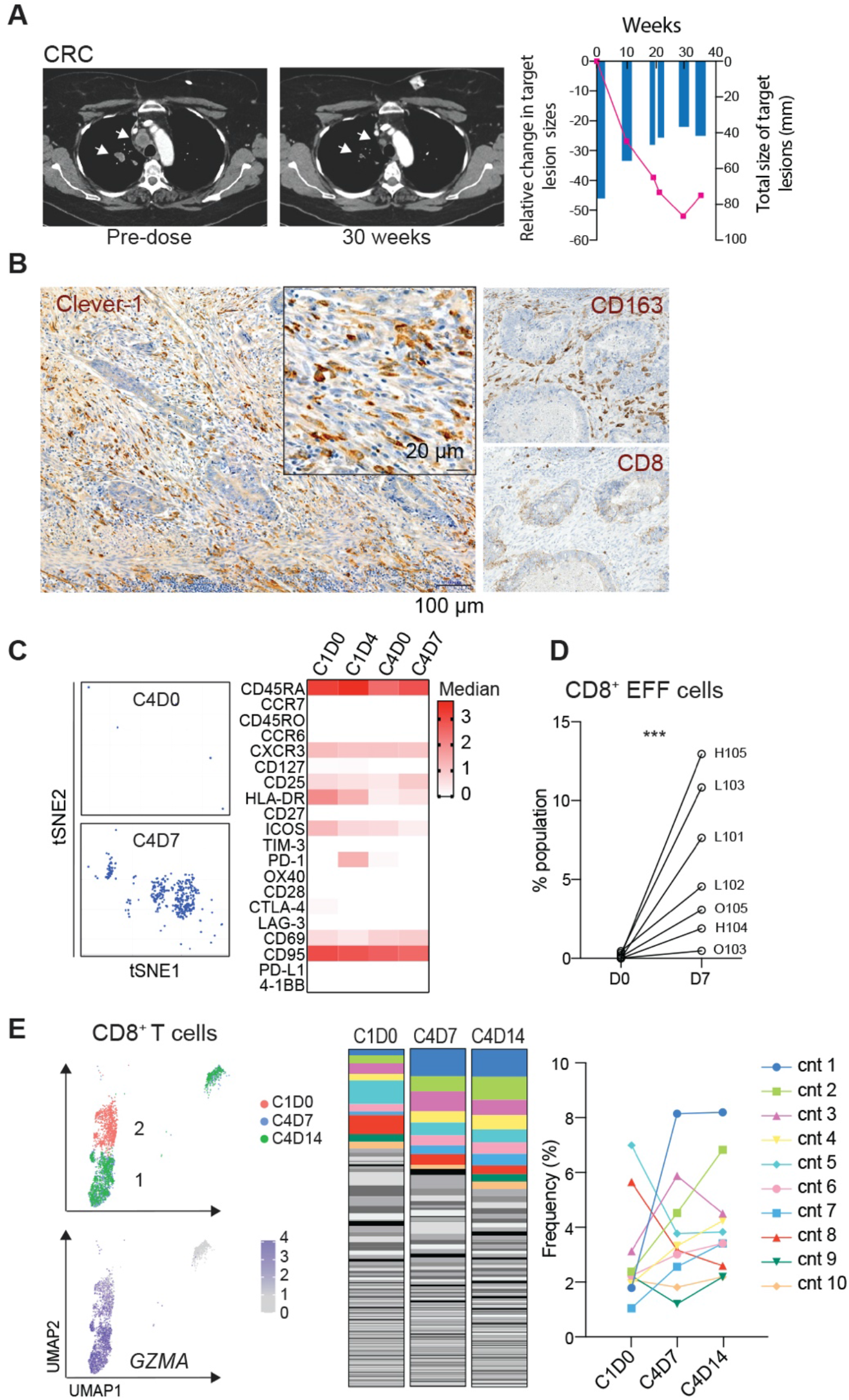
FP-1305-induced anti-tumor responses are reflected by clonal expansion of systemic effector CD8^+^ T cells. **A**, CT images of a patient (Pt6) with microsatellite stable (MSS) colorectal carcinoma (CRC) (six previous lines of therapy). Arrows highlight shrinking lung metastases. Size of the target lesions and change over time are presented on the right. **B**, Immunohistochemical analysis of Clever-1, CD163 and CD8 (brown) in archived rectal carcinoma tissue from the CRC patient. **C**, tSNE plot and median expression level of the cell surface markers of patient (Pt6) CD8^+^ EFF T-cell population at cycle 1 and 4 timepoints. **D**, Percent change of the CD8^+^ EFF population in patients 7 days after receiving FP-1305 in cycle 1. Student’s paired two-tailed *t*-test. **E**, Uniform Manifold Approximation and Projection (UMAP) plots of CD8 T-cell transcriptome colored by timepoint and expression of granzyme A (*GZMA*). Abundance of diverse CD8 T-cell clonotypes (cnt) at pre-dose (C1D0) and during cycle 4 (C4) timepoints. The ten most abundant TCR sequences are colored to show dynamics of specific clones during different timepoints. *** *p* <0.001

The appearance of the effector CD8 T-cell population prompted us to investigate these cells more thoroughly and identify whether these originated from clonal expansion of antigen experienced lymphocytes. Single-cell (sc)RNA-sequencing together with TCR sequencing of the responding patient’s circulating CD8^+^ T-cells showed clonal expansion of granzyme A (*GZMA*) high T-cell clonotypes (cnt) at cycle 4 (**Fig. 6E** and Supplementary Fig. 5A, B). These T-cells differed from pre-dose T-cells by higher expression of *FCGR3A* (CD16), *PFN1* (perforin), *DUSP1* (MKP-1) and *IL32* (**Fig. 6F** and Supplementary Fig. 5C). Both CD16 and MKP-1 have been shown to contribute to T-cell activation and cytotoxicity (Björkström et al., 2008; Zhang et al., 2009). These data indicate that FP-1305 is able to induce proliferation of antigen specific cytotoxic CD8^+^ T-cells. Moreover, the appearance of a *TCF7* (TCF1) high CD8^+^ T-cell population at cycle 4 (Supplementary Fig. 5A, B) suggests induction of a memory response with self-renewing potential (Kratchmarov et al., 2018). Unfortunately, we were not able to study whether the identified T-cell clonotypes were present in the tumor due to unavailable post-biopsy material.

### FP-1305 treatment leads to reduced numbers of Clever-1^+^ macrophages and changes in intra-tumoral CD8^+^ T-cells

To analyze whether the immune activation and increase in circulating lymphocytes was associated with immune infiltration into the tumor, we studied the number of CD8^+^ T-cells in pre- and post-biopsy tumor samples. Paired tumor samples were possible only in five patients who demonstrated progressive disease. In one patient (E104), a peri-tumoral CD8^+^ T-cell profile changed to intra-tumoral CD8^+^ T-cells nine weeks post-dose (**Fig. 7A**). In another patient (H105) the presence of peri- and/or intratumoral CD8^+^ T-cells was only very limited prior to FP-1305 and increased substantially post-dose. Clever-1 staining did not appear to change dramatically (moderate weakening of signal intensity) in post-samples but the number of Clever-1^+^ macrophages were reduced (**Fig. 7B**).

**Fig. 7.**
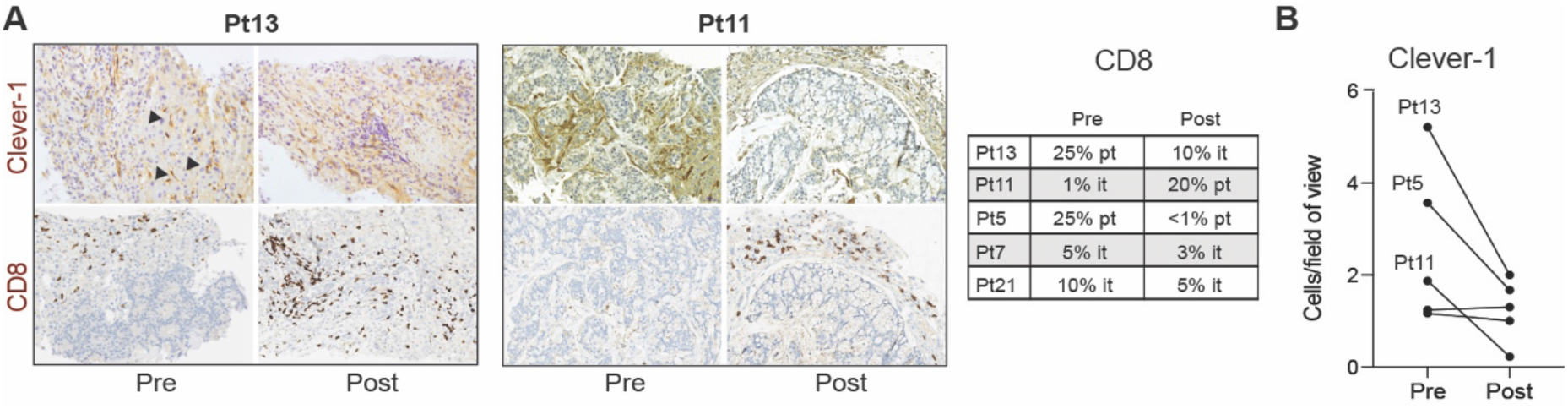
Post-treatment biopsy samples show reduced Clever-1 expression. **A**, Immunohistochemical staining of Clever-1 and CD8 in patient tumor biopsy samples pre- and post-treatment. Arrows point to Clever-1^+^ liver sinusoids. Quantification of peri and intra tumoral CD8 T-cells in biopsy samples. pt, peri tumoral; it, intra tumoral. **B**, Quantification of Clever-1^+^ macrophages in tumor biopsy samples pre- and post-treatment. Post-biopsy samples were taken after cycle 3.

## Discussion

The tumor microenvironment remains a major barrier to successful cancer eradication, limiting the efficacy of current checkpoint inhibitors. A range of pre-clinical studies confirm that macrophages are key orchestrators of this immunosuppressive niche, yet harnessing their therapeutic capability remains a significant challenge. Clever-1 is a macrophage scavenger receptor that is expressed on peripheral blood monocytes and is markedly upregulated by, for example, glucocorticoids on macrophage subsets supporting tissue tolerance (Kzhyshkowska et al., 2006; Palani et al., 2011). Macrophages express a range of scavenger receptors and historically there has been an assumption that there is significant redundancy within this large receptor family. Yet previous human *in vitro* and murine *in vivo* studies have highlighted the potential of inhibiting Clever-1 to skew macrophages to have a pro-inflammatory phenotype (Palani et al., 2016; Viitala et al., 2019).

The notable immunological finding from this early phase trial is that anti-Clever-1 treatment can induce robust peripheral T-cell activation in cancer patients with advanced disease. The systemic immune activation is a promising feature for clinical anti-tumor activity of FP-1305 since peripheral T-cell expansion has been reported to predict tumor infiltration and clinical response to anti-PD-L1 therapy (Wu et al., 2020). Moreover, recent findings indicate that newly generated transitory effector CD8^+^ T-cells emerging from lymphoid tissue resident stem-like CD8^+^ T-cells (TCF1^+^) play a critical role in PD-1-based immunotherapy (Hudson et al., 2019), a subset which was detectable after cycle 4 of FP-1305 therapy (Supplementary Fig. 5A). The downregulation of negative immune checkpoints on CD4^+^ T-cell subsets after one week on FP-1305 treatment further supports the induction of newly activated lymphocytes from the peripheral lymphoid organs. Where pre/post tumor biopsies were available, there were also features of increased CD8^+^ infiltration after treatment in selected cases, suggesting that FP-1305 could also have a direct impact on the tumor microenvironment.

Clever-1 contains multiple binding sites for various ligands (Kzhyshkowska et al., 2006) and it is not surprising that antibodies targeting non-overlapping epitopes on Clever-1 can have different functional readouts of known Clever-1 activities. Indeed, co-immunoprecipitation of Clever-1 with 9-11 identified an interaction of Clever-1 with several vacuolar ATPase complex proteins that was not identified with FP-1305. This clearly suggests that FP-1305 might out-compete v-ATPase complex binding and inhibit endosomal acidification. This seemed to be the case when cellular acidification was measured in primary human macrophages treated with FP-1305 but not 9-11. The recruitment of ATP6V0A1 to lysosomes is a critical component for lysosomal acidification and degradation of proteins. The duration of this process determines how well engulfed peptides can be loaded on MHC class I (cross-presentation) and presented on the cell surface to CD8^+^ T-cells. When we tested the ability of tumor-conditioned macrophages to cross-present engulfed ovalbumin, the cells genetically devoid of Clever-1 outperformed those of wildtype origin. This supports our clinical findings of the increased adaptive immune activation in patients receiving FP-1305.

The observation that FP-1305 can inhibit the scavenging of acLDL (Rantakari et al., 2016) suggests that this is the primary cause of the downregulation of the LXR/RXR and PPAR nuclear receptor pathways in MATINS patient monocytes. The LXR/RXR and PPAR receptors regulate lipid metabolism in monocytes (Nagy et al., 2012), and it is well established that these pathways shape the immune response of myeloid cells (Kidani and Bensinger, 2012). Their downregulation potentially drives the adaptive immune activation in heavily pre-treated patients. Recently, Donadon and colleagues identified a morphologically distinct tumor macrophage subset in colorectal liver metastases that associated with poor disease-free survival. The authors identify LXR/RXR as the most enriched pathway in these large macrophages, which was related to genes involved in cholesterol metabolism, scavenger receptors, MERTK and complement (Donadon et al., 2020). From a clinical point of view, indirectly regulating lipid metabolism by targeting Clever-1 may be beneficial because the actions are cell-specific and therefore prevent divergent responses or side effects from systemic targeting of these pathways.

In conclusion, therapeutic blockade of Clever-1 unveils a novel non-redundant pathway linking the innate and adaptive immune system in humans. Anti-scavenger receptor therapy therefore has the capability to convert immunologically ignorant tumors to an immune activated state. Our deep immunophenotyping of patient samples provides a strong scientific rationale to further explore the efficacy of Clever-1 as an anti-tumor target.

## Data Availability

Material requests should be addressed to Maija Hollmen. FP-1305 is a proprietary-owned antibody of Faron Pharmaceuticals. Requests for this should be addressed to Jami Mandelin.

## Author contributions

MH designed and directed the project with input from RV, JR, MV, SJ, MKK and JM. RV, JR, MV, TL, and MH carried out the experiments. RV, JR, MV, FV, AT, SJ, JM, MKK and MH analysed the data and interpreted results. JK, PJ, AP, SS, MJ, DR, YTM, TS, AM, and PB were involved in managing or executing the clinical trial at the study sites. MH wrote the manuscript with input from all authors.

## Acknowledgements

We thank Mari Parsama, Teija Kanasuo, Sari Mäki and Riikka Sjöroos for excellent technical assistance, and the Cell Imaging and Cytometry Core Facilities at Turku Bioscience Center for their help in mass cytometry. We also want to thank all the patients for participating in the clinical trial. A humble recognition should be addressed to Maria Lahtinen and Mari Kimpanpää for managing patient sample logistics at Faron Pharmaceuticals Ltd, and Maria Jokinen, Maria Oliveira and Jarna Hannukainen at Faron Pharmaceuticals Ltd and Laura Gardner at Simbec-Orion for clinical management of the MATINS study. The study was supported by Finnish Functional Genomics Centre, University of Turku, Åbo Akademi and Biocenter Finland. This study was funded by the Academy of Finland (AT, TL, SJ and MH), Emil Aaltonen Foundation (RV), Sigrid Jusélius Foundation (MH), and the Finnish Cancer Foundations (MH). Faron Pharmaceuticals sponsored the MATINS trial.

## Conflict of Interest

SJ, MKK, JM and MH own shares of Faron Pharmaceuticals. MH reports receiving funding from Faron for the preclinical development of anti-Clever-1 mAbs. SS, PB and MH have received consultancy fees from Faron. PB reports (outside the submitted work) honoraria from Bristol-Myers Squibb, MSD, Pfizer, Novartis, Oncorena, TILT Biotherapeutics, Ipsen, EUSA and Herantis Pharma, and stock ownership: TILT Biotherapeutics and Terveystalo.

Material requests should be addressed to Maija Hollmén. FP-1305 is a proprietary-owned antibody of Faron Pharmaceuticals. Requests for this should be addressed to Jami Mandelin.

## Methods

### Study design, patients and procedures

This multi-institutional, first in-human, open-label, non-randomized phase I, dose escalation study was approved by local institutional review boards. All participating patients signed informed consent. Patients in the Part I of the MATINS (NCT03733990) trial were dosed between 18 Dec 2018-28 Apr 2020 in the study sites at Helsinki, Oulu, and Tampere University Hospitals, Erasmus MC/Cancer Institute, Royal Marsden London and University Hospitals Birmingham. The data cut-off date for protocol-specified Part I analysis was May 7th, 2020. Participation was offered for subjects with advanced (inoperable or metastatic), treatment-refractory, histologically confirmed hepatobiliary, pancreatic, colorectal or ovarian cancer or cutaneous melanoma, without standard treatment options. Melanoma patients had to be immunotherapy-refractory (progression on or after PD-1 or CTLA-4 antibody therapy). Patients meeting the inclusion and none of the exclusion criteria specified in the study protocol (EudraCT Number: 2018-002732-24) were consented, pre-scanned (CT) for metastatic lesions and pre-biopsied before FP-1305 dosing.

Eligible patients had no cancer therapy for at least 3 weeks before first FP-1305 dosing. Patients were ≥18 years old with a life expectancy of > 12 weeks, an Eastern Cooperative Oncology Group (ECOG) performance status of 0 or 1, adequate organ function, and no ongoing systemic infections or active autoimmune disease. Concurrent antineoplastic therapies or systemic steroids were not permitted. Patients with measurable or non-measurable disease were allowed to be enrolled in the Part I.

### Sample randomization

The patient samples were analyzed as they arrived at Medicity in a blinded fashion not knowing patient clinical characteristics or drug responses. Since only 9 mL of blood was obtained for each patient at indicated timepoints, analyzes, including CyTOF, LPS/PMA stimulation and RNA sequencing, were not able to be performed uniformly on all patient samples. Thus, these were also randomly assigned per patient (Supplementary Table 1). A written informed consent was obtained from healthy donors donating cells to assess Clever-1 expression on blood monocytes. The samples were kept anonymous and handled according to the ethical guidelines set by the University of Turku.

### Sample collection and PBMC isolation

Heparin blood was collected at indicated time points at the clinical study sites. Blood was delivered overnight to Turku, Finland at room temperature. PBMCs were isolated with Ficoll-Paque density gradient centrifugation according the manufacturer’s instructions (GE Healthcare). Isolated cells were used fresh for flow cytometry and LPS induction assays and the remaining cells used for CyTOF and T-cell activation were frozen in RPMI medium (RPMI1640 (Sigma-Aldrich), 10% FCS and 2 mmol/L L-glutamine) supplemented with 10% DMSO for later use.

### CyTOF antibodies and antibody labelling

The used antibodies and the corresponding clone, the metal tag and provider are listed in Table 1. The antibodies labelled in-house were obtained as carrier free (CD45, CD197/CCR7, Ki67, CD169, CD8) or diluted (9-11 and FP-1305) in the protocol R-buffer 200 μg/mL and used 500 μL/labelling. Labelling was done with MaxPar labelling kits (MaxPar labelling kit, Fluidigm) following manufacturer’s instructions.

### CyTOF sample preparation

Frozen PBMCs were thawed and washed with RPMI medium. Cells were re-suspended in PBS and counted. 1-3×10^6^ cells per sample were stained with 2,5 μM Cell-ID cisplatin (Fluidigm; cat. 201064) viability reagent for 5 min at room temperature (RT). Thereafter the cells were barcoded with heavy-metal isotope-labelled anti-human CD45 (clone H130) antibodies (CD45_89Y (Fluidigm), CD45_141Pr (Fluidigm) and CD45_147Sm (BioLegend), 1/200) for 30 min at RT, washed carefully, combined and divided equally into two samples. The samples were blocked with human Kiovig solution (0.2 mg/ml) for 15 min at RT and stained with heavy-metal isotope-labelled anti-human antibody cocktails (PANEL 1 and 2) including the cell surface markers for 30 min at RT.

**PANEL 1.**

| Tag | Marker | Clone | Source |
| --- | --- | --- | --- |
| 154Sm | CD3 | UCHT1 | Fluidigm |
| 145Nd | CD4 | RPA-T4 | Fluidigm |
| 146Nd | CD8a | RPA-T8 | Fluidigm |
| 143Nd | CD127 | A019D5 | Fluidigm |
| 167Er | CD27 | O323 | Fluidigm |
| 165Ho | CD45RO | UCHL1 | Fluidigm |
| 169Tm | CD45RA | HI100 | Fluidigm |
| 170Er | HLA-DR | L243 | Fluidigm |
| 149Sm | CD25 | 2A3 | Fluidigm |
| 163Dy | CXCR3 | G025H7 | Fluidigm |
| 160Gd | CD28 | CD28.2 | Fluidigm |
| 176Yb | CD196/CCR6 | G034E3 | Fluidigm |
| 144Nd | CD69 | FN50 | Fluidigm |
| 155Gd | CD279/PD-1 | EH12.2H7 | Fluidigm |
| 159Tb | CD274/PD-L1 | 29E.2A3 | Fluidigm |
| 161Dy | CTLA-4 | 14D3 | Fluidigm |
| 158Gd | OX40 | ACT35 | Fluidigm |
| 175Lu | LAG-3 | 11C3C65 | Fluidigm |
| 153Eu | TIM-3 | F38-2E2 | Fluidigm |
| 173Yb | CD137/4-1BB | 4B4-1 | Fluidigm |
| 152Sm | CD95 | DX2 | Fluidigm |
| 168Er | CD278/ICOS | C398.4A | Fluidigm |
| 166Er | CD197/CCR7 | G043H7 | BioLegend |

**PANEL 2.** Grey denotes intracellular staining

| Tag | Marker | Clone | Source |
| --- | --- | --- | --- |
| 170Er | CD3 | L243 | Fluidigm |
| 144Nd | CD11b | ICRF44 | Fluidigm |
| 146Nd | CD64 | 10.1 | Fluidigm |
| 151Eu | CD14 | M5E | Fluidigm |
| 209Bi | CD16 | 3G8 | Fluidigm |
| 176Yb | CD56 | NCAM16.2 | Fluidigm |
| CD156Gd | CXCR6 | K041E5 | BioLegend |
| 165Ho | CD19 | HIB19 | Fluidigm |
| 162Dy | CD11c | Bu15 | Fluidigm |
| 173Yb | HLA-DR | L243 | Fluidigm |
| 150Nd | CD86 | IT2.2 | Fluidigm |
| 159Tb | CD274/PD-L1 | 29E.2A3 | Fluidigm |
| **154Sm** | **CD163** | GHI/61 | Fluidigm |
| **171Yb** | **CD68** | Y1/82A | Fluidigm |
| **168Er** | **CD206** | 15-2 | Fluidigm |
| **166Er** | **Clever-1** | 9-11 | InVivo |
| **158Gd** | **Clever-1** | CP12 | Antitope |

The samples stained with PANEL 1 were incubated with DNA intercalation reagent (1/1000, Cell ID Intercalator-103Rh in MaxPar® Fix and Perm Buffer; cat. 201067; Fluidigm) for 1h at RT, washed and fixed with 4% PFA solution overnight (o/n) at +4°C. The samples stained with PANEL 2 were fixed and permeabilized with the Transcription Factor Staining Buffer Set (00-5523-00; Invitrogen), blocked with human Kiovig solution (0.2mg/ml) for 15 min at RT and stained with heavy-metal isotope-labelled anti-human intracellular antibody cocktail for 30 min at RT. After washing, the samples were incubated with DNA intercalation reagent and washed and fixed as explained above.

Both panels were updated for the second set of staining with additional antibodies, which were: PANEL 1; Ki67_156Gd (clone Ki-67, Biolegend), PANEL 2; CD4_145Nd (clone RPA-T4), CD8_156Gd (clone SK1), Perforin_175Lu (clone PD48), Ki-67_161Dy (clone B56) and Cleaved caspase 3_142Nd (clone D3E9) all from Fluidigm. All the other added antibodies except CD4 and CD8 for PANEL 2 were intracellularly stained following the protocol described above.

The next day the samples were washed, resuspended in MaxPar Water (cat. 201069; Fluidigm) containing 1/10 dilution of EQ 4 Element Beads (Fluidigm) and immediately acquired by a CyTOF mass cytometer (Helios, Fluidigm). After bead normalization of the samples, viable singlet cells were de-barcoded by using FlowJo10 (TreeStar). In PANEL 1 CD45^+^CD3^+^ cells and in PANEL 2 CD45^+^CD3^-^ or CD45^+^ cells were gated and exported for further analysis.

### CyTOF data analysis

The data analysis was performed similarly as in Kimball et al 2018 J Immunol (Kimball et al., 2018). R studio version 1.2.1335 was downloaded from the official R Web site and Cytokit package was downloaded from Bioconductor and opened in R. Manually gated events (gated as explained above) were imported into Cytokit and subjected to Phenograph analysis. For PANEL 1, clustering was performed by using CD4, CD8, CD45RA, CCR7, CD45RO, CD127, CD25, CCR6, CXCR3 markers with additional settings: merge method; minimum, transformation; CytofAsinh, cluster method; Rphenograph, visualization method; tSNE and cellular progression NULL. For PANEL 2, clustering was performed by using CD3, CD11b, CD64, CD14, CD16, CD56, CD19, CD11c and HLA-DR markers with the same settings as above. Additionally, when running samples with updated panels also CD4 and CD8 markers were used in PANEL 2 clustering. Clusters were defined by Phenograph and these clusters were displayed on tSNE blots by using R package “shiny” to visualize different patients before and after treatment. Cluster colours, identification numbers, dot and label size were customised in “shiny”.

For each patient, the relative size of each cluster was calculated at both timepoints. A cluster x patient matrix was formed from the arithmetic differences in relative cluster sizes between the two timepoints. The matrix was then clustered using hierarchical clustering with Euclidean distance and complete linkage and plotted as a heatmap (R package ComplexHeatmap, function Heatmap) along with black filled circles representing relative cluster sizes at each timepoint, with circle radiuses proportional to the square root of the relative cluster sizes.

For the expression of each marker in each cell cluster, a median was calculated from the marker expressions of individual cells in that cluster. This calculation was done separately at both timepoints and for each patient. The change in marker expressions for each cluster between the two timepoints was assessed by applying a one-sample t-test on the differences of medians (R function t.test). P values were corrected for multiple testing using False Discovery Rate correction (Benjamini-Hochberg https://www.jstor.org/stable/2346101?seq=1, R function p.adjust). A marker x cluster matrix was formed from the FDR-corrected negative log10-transformed P values multiplied by the sign of the effect (positive or negative). The matrix was then clustered using hierarchical clustering with Euclidean distance and complete linkage and plotted as a heatmap (R package ComplexHeatmap, function Heatmap(Gu et al., 2016)). FDR-corrected P values smaller than 0.05 were considered significant.

### Flow cytometry

PBMCs were plated 0.2×10^6^ cells per well in a round-bottom 96-well plate (Sarstedt). All wells were stained with anti-human CD14-Pacific Blue (clone M5E2, BD Pharmingen) together with 10 μg/mL in-house conjugated anti-Clever-1 antibodies: 9-11 (InVivo Biotech) or FP-1305 (clone CP12, Abzena). Conjugation was performed using Alexa Fluor 647 Protein Labeling Kit from ThermoFisher according to manufacturer’s instructions. Irrelevant isotype control antibodies rat IgG2a-alexa647 (BD Pharmingen) or human in-house conjugated IgG4 (S241/L248E)-alexa647 (Antitope) were used for signal normalization, respectively. Fixed samples were run with LSRFortessa (BD) and analyzed with FlowJo10.

### RNA-sequencing

CD14^+^ monocytes were isolated from patient pre-dose (D0) and post-dose (D1, D7) heparin blood samples by Ficoll-Paque PLUS density-gradient centrifugation followed by magnetic enrichment with human CD14 microbeads (cat.130-050-201, Miltenyi Biotec) on MS Columns (cat. 130-042-201, Miltenyi Biotec) according to manufacturer’s instructions. FACS staining with anti-CD14 (clone M5E2, cat. 558121, BD Pharmingen) and anti-CD11b (clone ICRF44, cat. 301318, BioLegend) confirmed > 97% monocyte purity. Monocyte RNA was extracted using TRIsure (cat. BIO-38033, BioLine) according to the manufacturer’s protocol. RNA quality was confirmed using 2100 Bioanalyzer with RNA 6000 Pico kit (cat. G2939BA and 5067-1513, Agilent). cDNA library preparation and sequencing were performed by Novogene (Cambridge, UK) according to standard Novogene mRNA sequencing protocols on Illumina NovaSeq 6000 platform, where 150-bp paired-end reads were generated. Raw reads were filtered to remove reads with adaptor contamination, uncertain nucleotides (N >10%) or low-quality nucleotides (>50% bases with Q-score <20). Obtained high-quality reads were mapped to the human reference genome GRCh38 with HISAT2 (Kim et al., 2015). Transcript abundancy was quantified using HTSeq (Anders et al., 2015) and gene expression levels calculated as FPKM (fragments per kilobase million). For hierarchical clustering and principal component analysis (PCA), raw read counts were regularized logarithm transformed with DESeq2 (Love et al., 2014). After transformation, genes with FPKM value >1 were selected for analysis. Unsupervised hierarchical clustering was performed based on Euclidean distances using complete linkage method. R version 3.6.2 (https://www.R-project.org/) was used for PCA and hierarchical clustering and figures were produced with packages pheatmap and ggplot2. For differential expression analysis, read counts were TMM (trimmed mean of M-values) normalized in edgeR(Robinson and Oshlack, 2010). Differently expressed genes in D1 and D7 samples were identified in each patient with DEGseq (Wang et al., 2010) as genes with Benjamini-Hochberg adjusted p-value <0.005 and |log_2_(FoldChange)| >1. Functional enrichment among differently expressed genes with FPKM value >1 was determined by performing canonical pathway analysis in Ingenuity Pathway Analysis software (IPA, Qiagen)(Krämer et al., 2014). The analysis was limited to genes and relationships described in monocytes or macrophages and p-value <0.05 was considered significant. The RNAseq data has been submitted to the Human Gene Omnibus under the accession code: GSE152169

### Quantitative (q)PCR

Total RNA was isolated from patient pre-dose (D0) and post-dose (D1, D7) CD14^+^ monocytes as described for RNAseq. Extracted RNA (100–300 ng) was reverse transcribed into cDNA using SuperScript VILO cDNA synthesis kit (cat. 11754-250, Thermo Fisher Scientific) according to manufacturer’s instructions. Quantitative PCR was performed by using pre-designed TaqMan gene expression assays (Applied Biosystems) for IL1R2 (Hs00174759_m1), JUN (Hs01103582_s1), PPBP (Hs00234077_m1), TNFRSF1A (Hs01042313_m1) and GAPDH (Hs02758991_g1) and TaqMan universal master mix II (cat. 440040, Applied Biosystems) following manufacturer’s instructions. Triplicate reactions (10 ul) with 4 ng of cDNA per well were made. The reactions were run using QuantStudio 3 Real-Time PCR system (Applied Biosystems) and QuantStudio design and analysis software v1.5.1. Relative quantification (RQ) values were calculated with the ΔΔCt method using GAPDH as an endogenous control.

### Human macrophage differentiation and polarization

Monocytes were positively enriched from PBMCs collected from buffy coats from healthy donors with CD14 Microbeads on LS-columns (130-042-401, Miltenyi Biotec). Monocytes were seeded to cell culture dishes at 1 × 10^6^ cells/ml in IMDM (31980022; Thermo Fisher Scientific) supplemented with 10 % FBS (F7524; Merck), penicillin/streptomycin (15140-122; Thermo Fisher Scientific) and 50 ng/ml recombinant human M-CSF (574806; Biolegend) and incubated for seven days at +37 °C and 5 % CO_2_ with a medium change on day 4. On day seven, the medium was changed to IMDM supplemented with 10 % FBS, penicillin/streptomycin, 100 nM dexamethasone (D-2915; Merck) and 20 ng/ml of human recombinant IL-4 (200-04-50UG; Peprotech) to induce M2 polarization for two days.

### KG-1 cell culture and differentiation

The Clever-1^high^ acute myelogenous leukemia cell line KG-1 was purchased from ATCC (CCL-246) and cultured in IMDM supplemented with 20 % FBS and penicillin/streptomycin as recommended. To induce KG-1 macrophage differentiation, phorbol 12-myristate 13-acetate (PMA) (P8139; Merck) was added to 300 nM and the cells were incubated for another three days.

### Coimmunoprecipitation

M2 macrophages were detached by washing once with PBS and incubating for 30 min in 5 mM EDTA in PBS at room temperature followed by gentle scraping. The collected cells were lysed for 30 min in IP lysis buffer (20 mM Tris-HCl [pH 8.0], 137 mM NaCl, 1 % Triton X-100, 2 mM EDTA) supplemented with 2× cOmplete Protease Inhibitor Cocktail (11697498001; Merck) with rotation at +4 °C. The lysates were cleared with a 10 min centrifugation at 20,000 *g* and +4 °C and the supernatants were transferred to fresh tubes. The lysate was precleared and diluted to 3–5 mg/ml. Aliquots of 1 ml were incubated with 10 μg of AK FUMM 9-11 or FP-1305 antibody (both InVivo) or their isotype-matched irrelevant isotype-matched control antibodies rat IgG2a (clone eBR2a; 14-4321-82; Thermo Fisher Scientific) and human IgG4 (clone QA16A15; 403702; Biolegend), respectively, overnight with rotation at +4 °C. Then, the lysates were transferred to fresh tubes with 200 μl of Dynabeads Protein G beads (10003D; Thermo Fisher Scientific) and incubated for one hour with rotation at +4 °C. The beads were washed 3 × 5 min with 1 ml of IP lysis buffer supplemented with 1× cOmplete Protease Inhibitor Cocktail at +4 °C. After the final wash, the wash buffer was removed completely and the beads resuspended in 100 μl of 2× non-reducing SDS sample buffer (120 mM Tris-HCl [pH 6.8], 20 % glycerol, 4 % SDS). Immunoprecipitated proteins were eluted for 10 min with mixing at room temperature, after which the beads were removed and the eluates transferred to fresh tubes. For mass spectrometry samples, dithiothreitol was added to 50 mM, after which the samples were heated at +95 °C for 5 min and run approximately 1.5 cm into 10 % hand-cast SDS-PAGE resolving gels. The gels were stained with Coomassie Blue and the protein lanes were cut and submitted for mass spectrometric analysis.

### Mass spectrometric analysis

Mass spectrometric analysis was performed at the Turku Proteomics Facility, University of Turku and Åbo Akademi University. The samples were in-gel digested according to a standard protocol at the Proteomics Facility. Digested peptides were dissolved in 15 μl of 0.1 % formic acid and a suitable amount of peptides from each sample was submitted to LC-ESI-MS/MS analysis. Washes and a blank run were submitted between samples to reduce carryover of highly abundant peptides. The LC-ESI-MS/MS analysis was performed on a nanoflow HPLC system (Easy-nLC1200; Thermo Fisher Scientific) coupled to the Q Exactive HF mass spectrometer (Thermo Fisher Scientific) equipped with a nano-electrospray ionization source. Peptides were first loaded on a trapping column and subsequently separated inline on a 15 cm C18 column (75 μm × 15 cm ReproSil-Pur 5 μm 200 Å C18-AQ; Dr. Maisch HPLC GmbH). The mobile phase consisted of water with 0.1 % formic acid (solvent A) or acetonitrile/water (80:20 (v/v)) with 0.1 % formic acid (solvent B). A 30 or 60 min gradient from 8 to 43 % solvent B followed by a wash stage with 100 % solvent B was used to elute peptides.

### LC-MS/MS data acquisition and analysis

Mass spectrometric data was acquired automatically with Thermo Xcalibur 4.1 software (Thermo Fisher Scientific). An information-dependent acquisition method consisted of an Orbitrap MS survey of the mass range 300–2000 *m*/*z* followed by HCD fragmentation. Data files were searched for protein identification using Proteome Discoverer 2.3 software (Thermo Fisher Scientific) connected to an in-house server running the Mascot 2.6.1 software (Matrix Science). Data were searched against the SwissProt database with the taxonomy filter *Homo sapiens*, trypsin as the enzyme, oxidation and acetylation as variable modifications, carbamidomethylation as a fixed modification, ±10 ppm as the peptide mass tolerance, ±0.02 Da as the fragment mass tolerance, maximum missed cleavages as 2 and the instrument type as ESI-TRAP. A minimum of two peptides per protein were used to filter the results. The criteria used for including a protein as a 9-11- or FP-1305-specific hit were that the protein was present in at least two out of three biological replicates, the protein was identified by at least three unique peptides and that the protein was not identified in the IgG control or it had an over 3-fold enrichment of uniquely identified peptides over the IgG control. To detect likely contaminants among the mass spectrometry hits, we uploaded the spectral count data of the 9-11-and FP-1305-specific proteins into CRAPome (Mellacheruvu et al., 2013) and analyzed them against 16 selected CRAPome controls (cell type HEK293, total cell lysate, Dynabeads magnetic affinity support) using default settings. FC-A > 4 was used as the cut-off value to exclude the most commonly found contaminants, which consisted mostly of nuclear and ribosomal proteins and ribonucleoproteins. Reported high-confidence (>0.7) protein–protein interactions between the hit proteins were downloaded from the STRING database (Szklarczyk et al., 2017) and visualized with Cytoscape 3.7.2 (Shannon et al., 2003). The yFiles Organic layout was applied and clusters identified with the Markov Clustering (MCL) algorithm of the clusterMaker app using default settings (Morris et al., 2011). Gene Ontology Biological Process (GO BP) terms enriched in the five largest clusters were identified with the STRING Enrichment app (redundancy cut-off 0.7) (Doncheva et al., 2019). For Ingenuity Pathway Analysis (IPA), lists of proteins specific for 9-11, specific for FP-1305 or shared by both were uploaded to IPA and submitted for core analysis.

### Clever-1 RNA interference and KG-1 macrophage differentiation

The day before transfection, KG-1 cells were replenished 1:1 with fresh medium. On the day of transfection, KG-1 cells were counted, washed three times with PBS to remove serum and re-suspended to 20 × 10^6^ cells/ml in OPTI-MEM reduced serum medium (51985026; Thermo Fisher Scientific) at room temperature. The suspension was split into 100 μl aliquots per electroporation and siRNA dissolved in RNase-free water was added to 2 μM. The siRNAs used in this study were: pooled scramble control siRNA (ON-TARGETplus Control Pool D-001810-10-20; Dharmacon) and single siRNAs targeting the human Clever-1 RNA sequences AUGAUGAGCUCACGUAUAA (siR1) or UCAAGUCGCUGCCUGCAUA (siR2) (J-014103-05-0020 and J-014103-08-0020, respectively; Dharmacon). The mixture of KG-1 cells and siRNA was transferred to electroporation cuvettes (P45050; Thermo Fisher Scientific), electroporated with program U-001 on the Nucleofector II device (Amaxa Biosystems) and transferred to pre-warmed IMDM supplemented with 20 % FBS and penicillin/streptomycin for overnight recovery and differentiated with PMA as described. Cells remaining in suspension were removed by gentle PBS washes and the adherent KG-1 macrophages were used for experiments. Clever-1 knockdown was verified by surface staining followed by FACS analysis.

### Endocytosis and acidification experiments

To determine total cellular pH, LysoSensor Green DND-189 (L7535; Thermo Fisher Scientific) was added 1:1,000 to pre-warmed culture medium with approximately 1 × 10^6^ cells/ml, after which the cells were incubated for the indicated time points before proceeding with FACS analysis. To analyze DQ-OVA antigen degradation (D12053; Thermo Fisher Scientific) and acLDL uptake and acidification, the cells were re-suspended at approximately 1 × 10^6^ cells/ml in room-temperature CO_2_-independent medium (18045054; Thermo Fisher Scientific) supplemented with L-glutamine (35050-038; Thermo Fisher Scientific) and penicillin/streptomycin and the indicated treatments added to 10 μg/ml. The cells were incubated at +16 °C for 30 min to allow initial surface binding, washed three times with ice-cold PBS, re-suspended in pre-warmed culture medium and incubated for the indicated time points before proceeding with FACS analysis. Native human acetylated low-density lipoprotein (acLDL) was purchased from Bio-Rad (5685-3404) or Thermo Fisher Scientific (L35354) and labelled with the Alexa Fluor 488 or 647 Microscale Protein Labeling Kit (A30006 and A20173, respectively; Thermo Fisher Scientific), the pHrodo iFL Green Microscale Protein Labeling Kit (P36015; Thermo Fisher Scientific) according to the manufacturer’s instructions. Before FACS analysis, the cells were washed three times with 0.1 % BSA in PBS and finally re-suspended in PBS. To quench extracellular fluorescence, trypan blue (15250061; Thermo Fisher Scientific) was added to a 0.1 % final concentration immediately before FACS analysis. Standard curves for cells treated with LysoSensor Green or acLDL labeled with pHrodo Green were prepared with the Intracellular pH Calibration Buffer Kit (P35379; Thermo Fisher Scientific) according to the manufacturer’s instructions.

### Antigen presentation

For murine macrophage differentiation, bone marrows from age- and sex-matched wildtype and Clever-1 knockout mice were collected by flushing with MACS buffer (PBS supplemented with 0.5 % BSA and 2 mM EDTA) (Karikoski et al., 2014). Collection of mouse bone marrow was performed in adherence to the Finnish Act on Animal Experimentation (62/2006) and was approved by the Committee for Animal Experimentation (license number 5762/04.10.07/2017). Red blood cells were lysed with PharmLyse according to the manufacturer’s instructions (555899; BD). Bone marrow cells were seeded to cell culture dishes at 1 × 10^6^ cells/ml in IMDM (31980022; Thermo Fisher Scientific) supplemented with 10 % FBS (F7524; Merck), penicillin/streptomycin (15140-122; Thermo Fisher Scientific) and 50 ng/ml recombinant murine M-CSF (576406; Biolegend) and incubated for seven days at +37 °C and 5 % CO_2_ with a medium change on the fourth day. On day seven, the medium was removed and the adherent macrophages were washed once with PBS. Macrophages were detached by incubating for 30 min in 5 mM EDTA in PBS at room temperature followed by gentle scraping. The collected cells were TAM-polarized for 48 h in low-adherence U-bottom 96-well plates (7007; Costar) at 1 × 10^6^ cells/ml in IMDM supplemented with 50 % sterile-filtered LLC1-conditioned medium (collected from LLC1 cell cultures three days after subculturing when flasks were ∼70 % confluent, passed through 0.22 μm filters [SLGS033SS; Millipore] and stored at −20 °C). The following treatments were added to polarized macrophages for 6 h before FACS analysis: ovalbumin SIINFEKL peptide at 10 ng/ml (vac-sin; InVivoGen), full-length ovalbumin protein at 1 mg/ml (LS003059; Worthington) or UV-irradiated apoptotic ovalbumin-expressing EG.7 cells (ATCC CRL-2113) 1:1 with macrophages. The macrophages were stained with fluorochrome-conjugated antibodies against F4/80 (12-5743-82; Invitrogen), Clever-1 (described below) and the SIINFEKL/MHC I complex (11-4801-85; Invitrogen) and analyzed as described below.

### Western blot analysis

For western blot analysis, appropriate amounts of protein were diluted to 1× SDS-PAGE sample buffer (40 mM Tris-HCl [pH 6.8], 1.0 % SDS, 5 % glycerol, 0.0003 % bromophenol blue, 50 mM DTT) and heated at +95 °C for 5 min. For detecting Clever-1, DTT was omitted and the samples heated at +70 °C for 10 min. Protein samples were resolved by electrophoresis on handcast SDS-PAGE gels and transferred to PVDF membranes (1704156; Bio-Rad) using the Trans-Blot Turbo standard program (Bio-Rad). Membranes were blocked with 5 % nonfat dry milk in Tris-buffered saline (10 mM Tris-HCl [pH 7.6], 150 mM NaCl) supplemented with 0.1 % Tween-20 (TBST) and then western blotted by standard methods. Primary antibodies used for western blotting were: anti-ATP6V0A1 (NBP1-59949; Novus Biologicals), anti-ATP6V1A (PA5-29191; Invitrogen), anti-TCIRG1 (PA5-90425; Invitrogen) and anti-Clever-1 (clone 3-372), all diluted 1:1,000. The blots were probed with HRP-conjugated secondary antibodies against mouse or rabbit (G21040, G21234; Invitrogen), diluted 1:5,000, and detected with Pierce ECL western blotting substrate (32209; Thermo Fisher) on CL-XPosure films (34090; Thermo Fisher). Scanned images have been cropped for presentation.

### T-cell activation assay

PBMCs were plated 1-3×10^6^ cells per well (depending on the patients’ PBMC count) in ultra-low attachment 96-well plates (Corning) in IMDM (Gibco cat:21980-032) supplemented 1/1000 Brefeldin (BioLegend), 50 ng/mL phorbol 12-myristate 13-acetate (PMA) and 0.5 μg/mL ionomycin (both from Sigma) and incubated at 37°C in a 5% CO_2_ humidified chamber for 4 hours. The cells were washed, stained with Zombie Red viability dye (BioLegend) and cell surface stained with anti-human CD8-FITC (clone RPA-T8, BD Biosciences). The cells were fixed with 4% PFA and permeabilized using 0.3% Triton in PBS for 20 min RT and thereafter stained with anti-human IFNγ-APC (clone B27, BD Biosciences) and IL-2-BV421 (clone MQ1-17H12, BioLegend).

### Immunofluorescence

For anti-Clever-1 antibody internalization analysis, CD14^+^ monocytes were isolated as described above from EDTA-blood samples collected from healthy volunteer donors and frozen in HEC medium supplemented with 10% DMSO. Thawed monocytes (1×10^5^ per well) were differentiated into macrophages on 8-well ibiTreat chamber (cat. 80826, ibidi®) for five days and M2-polarized as described above. Anti-Clever-1 antibodies FP-1305 (clone CP12, Abzena) and 9-11 (InVivo Biotech), conjugated in-house to Alexa Fluor 647 (cat. A20173, Invitrogen) and Alexa Fluor 488 (cat. A10235, Invitrogen), respectively, or their conjugated isotype controls human IgG4 (S241/L248E, Antitope) and rat IgG2a (clone MEL-14, AK1632/01.2, InVivo Biotech) were added in the culture medium to 20 μg/ml for 5 min or 2h. The cells were then washed with cold PBS, fixed in 4% PFA for 15 min at RT and permeabilized in 0,1% triton X-100 for 15 min at RT. Nuclei were labeled with 4 μM Hoechst 33342 (cat. 62249, ThermoFisher) for 10 minutes. From the 5 min time point, z-stacks of the cells were captured with Marianas spinning disk, as described below. From the 2h time point, z-stacks of Clever-1+ cells were captured with an LSM 880 confocal microscope with an Airyscan detector (Carl Zeiss) and a 63x objective (oil, NA 1.4, C Plan-Apochromat, Carl Zeiss). Images were acquired and processed with Zen software (2.3 SP1 black edition, Carl Zeiss) and analyzed for co-localization with NIH ImageJ software (version 1.52p). After background subtraction with median filtering, Colocalization Threshold plug-in was used to calculate Manders’ co-localization coefficients M1 and M2 above Costes threshold (zero-zero pixels excluded) within a region of interest (ROI) spanning the whole cell area.

To assess Clever-1 and ATP6V0A1 co-localization after acLDL treatment, KG-1 cells (1×10^5^) were seeded on 8-well ibiTreat chamber and differentiated with PMA as described above. After three days, the adherent cells were treated with 10 μg/ml human acLDL conjugated to Alexa Fluor 647 for 3h. The samples were washed with cold PBS, fixed in 4% PFA for 15 min at RT and permeabilized and blocked in 0.1% triton X-100, 5 % goat serum (005-000-121, Jackson ImmunoResearch) and 0.1 mg/ml Kiovig (LE-072213, Baxter) in PBS for 20 min at RT. The cells were stained for Lamp1 (1:100, clone D2D11, cat. 9091, Cell Signaling), ATP6V0A1 (1:100, polyclonal, cat. H00000535-A01, Abnova) and Clever-1 (10 μg/ml, 9-11, InVivo Biotech) or control stained for rabbit IgG (2 μg/ml, polyclonal, cat. BE0095, BioxCell), mouse IgG (5 μg/ml, polyclonal, cat. 010-0102-0005, Rockland) and rat IgG2a (10 μg/ml, clone 2A3, #BP0089, InVivoPlus,) for 1h at RT. Anti-rabbit, anti-mouse and anti-rat secondary antibodies (1:300, cat. A31556, A11030 and A11006, ThermoFisher) were incubated for 30 min at RT. Z-stacks were acquired with Marianas spinning disk (Intelligent Imaging Innovations) connected to CSU-W1 scanning unit (Yokogawa) and Orca Flash 4 sCMOS camera (Hamatsu) using a 63x objective (oil, NA 1.4, Plan-Apochromat, Carl Zeiss). SlideBook 6.1 software (Intelligent Imaging Innovations) was used in image acquisition. Images were analyzed with ImageJ. Single stained samples were used to confirm the absence of bleed-through signal. Clever-1 and ATP6V0A1 intensity profiles across lines drawn over Clever-1+ vesicles were generated from background subtracted images using Plot Profile function. The amount of ATP6V0A1 in the lysosomes of KG-1 cells was quantified from z-stack mid-slices of 10-15 Clever-1+ cells for each staining. The areas of Lamp1+ vesicles within the cell were determined by applying a common threshold to the Lamp1 images and the quantity of ATP6V0A1 within these Lamp1+ vesicle ROIs was measured as mean fluorescence intensity and integrated density (IntDen) from background subtracted ATP6V0A1 images. ATP6V0A1 IntDen was normalized to cytoplasm area.

### IFNγ and CXCL10 measurement

Blood (2.5 ml) was collected to serum separation tubes (BD Vacutainer®) and clotted for 30 minutes at room temperature. Serum was separated with centrifugation, collected and stored below -70 °C. Samples were analyzed using V-PLEX Proinflammatory Panel 1 (Meso Scale Diagnostics) according to the kit instructions.

### LPS induction assay

PBMCs (1×10^5^) were plated in ultra-low attachment 96-well plates and incubated with or without TLR4 specific LPS (20ng/ml; LPS-EB Ultrapure, Invivo Gen) in IMDM for 24h. Samples were collected the next day and centrifuged 4000 rpm to remove the cells. The supernatant was frozen at – 70°C prior to analysis with Bio-Plex Pro™ Human Inflammation Panel 1, 37-Plex (#171AL001M, BioRad) according to the manufacturer’s protocol.

### Immunohistochemistry

Formalin-fixed paraffin-embedded (FFPE) tissue was stained with Vectastain Elite ABC-HRP rat kit (PK-6104, Vector Laboratories). Briefly, sections were deparaffinised and antigen retrieval for Clever-1 staining was done with proteinase K (S3020, Dako). Endogenous peroxidase removal was done with 1% H_2_O_2_-MQ solution in RT for 20 minutes followed by blocking for 1 hour. Anti-Clever-1 (Rat anti-human Clever-1 (clone S2-7/H7) hybridoma medium, in-house) and irrelevant rat IgG2a (clone MEL-14, AK1632/01.2, InVivo Biotech) isotype control staining was done o/n at +4°C. Anti-rat secondary antibody was used according to manufacturer’s instructions. ABC-reagent mixture was incubated for 30 minutes at RT and DAB Chromogen (K3468, Dako) was added according to the manufacturer’s instructions and incubated for 5 minutes at RT. Counterstaining was done in Mayer’s hematoxylin for 30 seconds followed by dehydration and mounting with DPX mounting medium (06522, Sigma). Slides were imaged with Panoramic 250 slide scanner (3DHISTEC). CD8 and CD163 staining was performed at Covance according to their validated protocols.

### Sc-RNA-seq with TCR sequencing

T-cells from cycles 1 and 4 of the CRC patient were enriched using CD3 microbeads (Miltenyi Biotec). Single cells were partitioned using a Chromium Controller (10× Genomics) and scRNA-seq and TCRαβ-libraries were prepared using Chromium Next GEM Single Cell 5′ Library & Gel Bead Kit (10× Genomics), as per manufacturer’s instructions (CG000207 Rev D). In brief, approximately 16,500 cells from each sample, suspended in 0.04% BSA in PBS were loaded on the Chromium Next GEM Single Cell G Chip, aiming the capture of 10,000 single cells. During the run, single-cell barcoded cDNA is generated in nanodroplet partitions. The droplets are subsequently reversed, and the remaining steps are performed in bulk. Full length cDNA was amplified using 14 cycles of PCR (Veriti, Applied Biosystems). TCR cDNA was further amplified in a hemi-nested PCR reaction using Chromium Single Cell Human T Cell V(D)J Enrichment Kit (10× Genomics). Finally, the total cDNA and the TCR-enriched cDNA was subjected to fragmentation, end repair and A-tailing, adapter ligation, and sample index PCR (14 and 9 cycles, respectively). The libraries were sequenced using an Illumina NovaSeq, S1 flowcell with the following read length configuration: Read1 = 26, *i*7 = 8, *i*5 = 0, Read2 = 91. The raw data was processed using Cell Ranger 3.1.0. with GRCh38 as the reference genome.

### Sc-RNA-seq data analysis

Post-processing including demultiplexing, read alignment and quality control were performed at the Institute for Molecular Medicine Finland (FIMM) using the 10X Genomics Cell Ranger package (v3.1.0). The Cell Ranger outputs were analyzed by Seurat (v3.1) for graph-based clustering, analysis of differentially expressed genes and extraction of TCR sequences from cell subsets. TCR clonotype data including clonotype ID and CDR3 amino acid sequences were added to the metadata for each cell, and data of C1D1, C4D8 and C4D15 were merged. For quality control genes, which were expressed in less than 5 cells were removed. We also filtered out cells that had less than 200 genes and cells with greater than 4000 genes and 10% mitochondrial genes. Data were normalized by the function of NormalizeData, and variable genes across the single cells were detected by the FindVariableFeatures. A linear transformation was applied on the data by the function of ScaleData prior to dimensional reduction. Then, FindNeighbors and FindClusters were used to apply nearest neighbor graph-based clustering to the cells using 15 dimensions and 0.6 resolution. The clustering was visualized with Uniform Manifold Approximation and Projection (UMAP) using RunUMAP with 15 dimensions. To analyze CD8 T-cell subsets and the clonality during the treatment, *CD8A* expressing CD8^+^ T-cell clusters were selected from initial clusters and subclustered by reapplying FindVariableFeatures, ScaleData, FindNeighbors, FindClusters and RunUMAP with 15 dimensions and resolution 0.6. Metadata of each CD8 T-cell were extracted and frequencies of each clonotype at each timepoint were calculated. The change in CD8 T-cell clones during the treatment were chased by analyzing the CDR3 amino acid sequences. The sc-RNA seq data has been submitted to the Human Gene Omnibus under the accession code: GSE152169.

### Statistics

Data are presented as mean ±SD unless otherwise noted with violin plots additionally showing individual data points. Comparisons between repeated measures were performed with One-way ANOVA (parametric), Friedman test (non-parametric) or Two-way ANOVA with Sidak’s multiple comparisons test. Group analysis between two timepoints (treatments, genotypes) was performed with two-sided (un)paired student’s *t*-test (parametric) or Mann-Whitney U test and Wilcoxon matched-pairs signed rank test (non-parametric). *P* < 0.05 was considered statistically significant. Statistical analyses were performed with Prism 8 (GraphPad).

---

**Supplementary Fig. 1.**
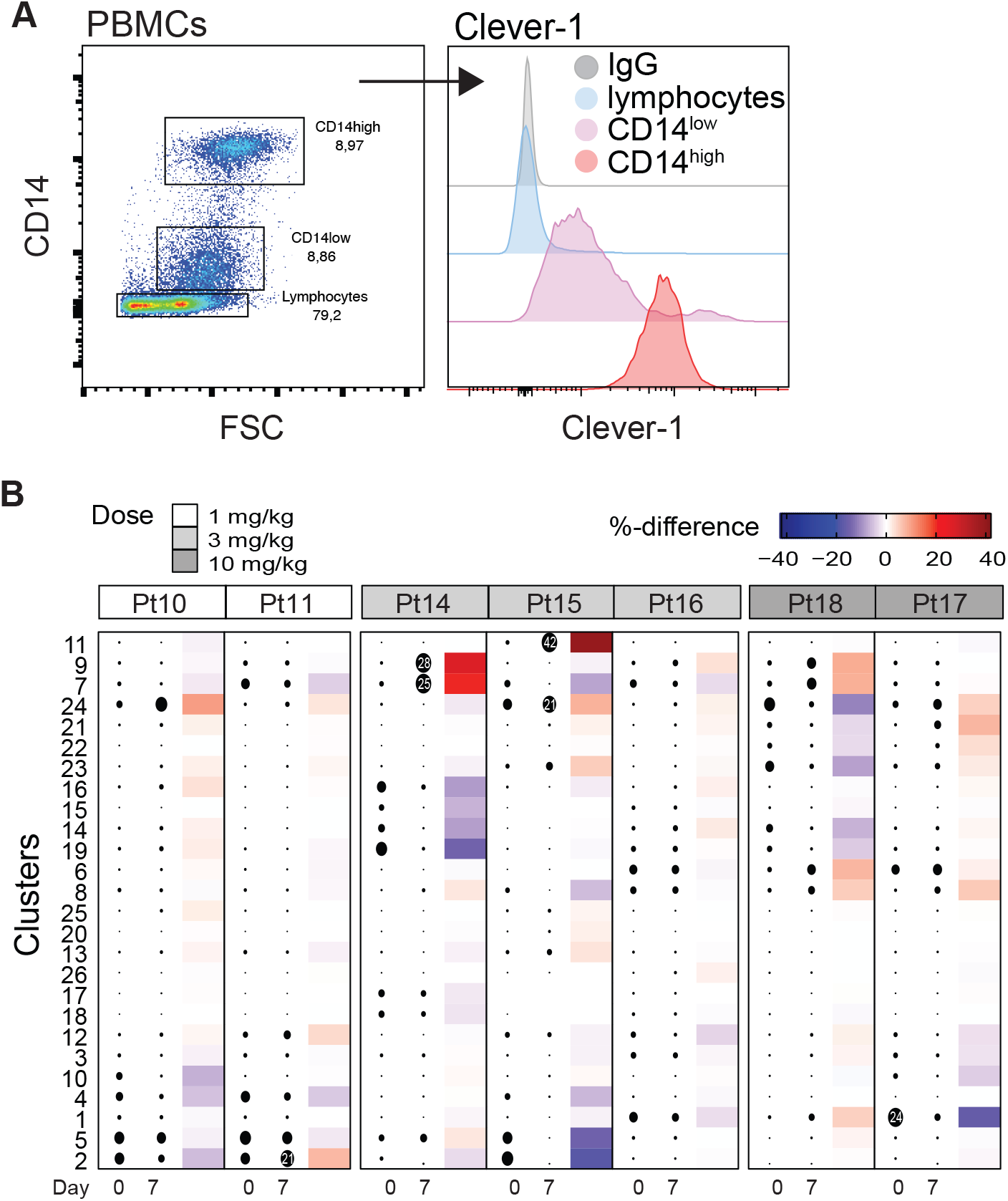
**A**, Flow cytometry analysis of cell surface Clever-1 expression on Ficoll-purified blood mononuclear cell populations (PBMC). **B**, Heatmap of cluster size pre- (D0) and post- (D7) treatment. Dot size indicates the relative percent of the cluster per sample. Red color points to positive and blue to negative change between pre- and post-samples within a patient (Related to Fig. 1A).

**Supplementary Fig 2.**
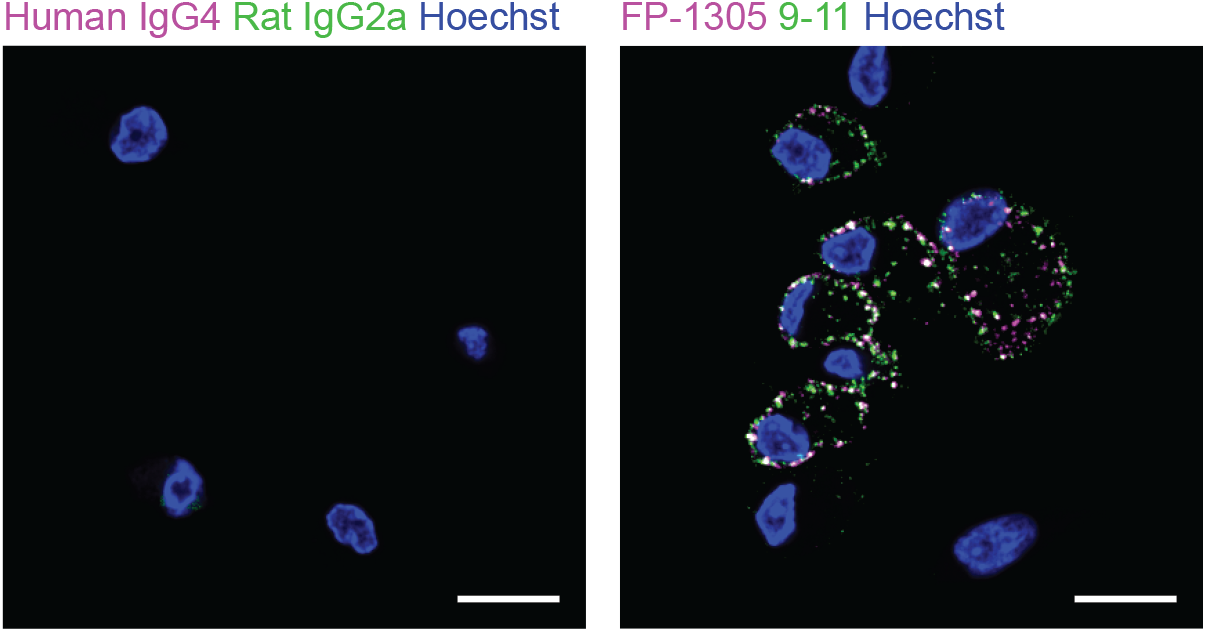
Confocal images of the uptake of FP-1305(AlexaFluor647) and 9-11(AlexaFluor488) by human dexamethasone polarized macrophages after 5 min. Human IgG4 and Rat IgG2a served as isotype controls for FP-1305 and 9-11, respectively. Images obtained using spinning disk, 190515. Scale bars 15 μm.

**Supplementary Fig. 3.**
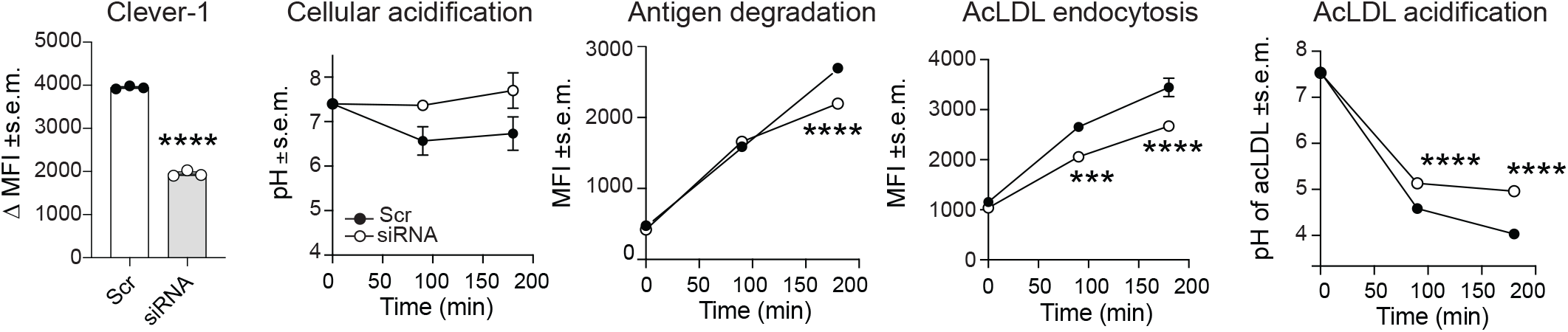
Clever-1 knockdown efficiency in KG-1 macrophages quantified by flow cytometry with 9-11 antibody. Total cellular acidification, antigen degradation, acLDL endocytosis and acLDL acidification kinetics was measured in KG-1 macrophages transfected with scramble (Src) or Clever-1 siRNA, all n=3. * p <0.05, ** p < 0.01, *** p < 0.001, **** p < 0.0001

**Supplementary Fig. 4.**
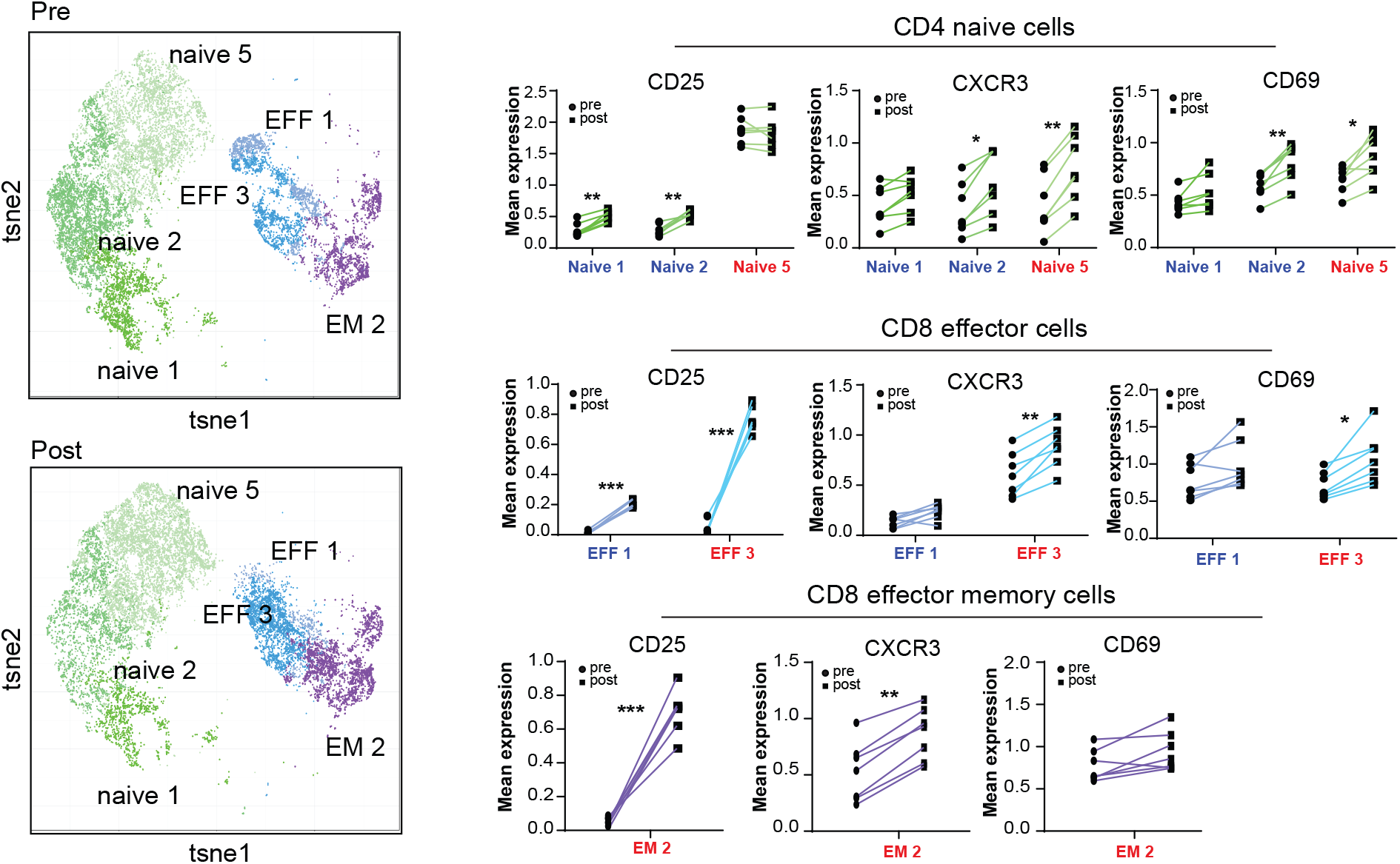
tSNE plots and mean expression of activation markers on T-cell clusters at pre-dose (D0) and day 7 (post). The color code blue on the x-axis title indicates that this cell population decreased (%) from pre- to post-dose, and the color red indicates an increase in this population. The %-changes relate to Fig. 2C. Paired student’s t-test. P *<0.05, **<0.01, ***<0.001.

**Supplementary Fig. 5.**
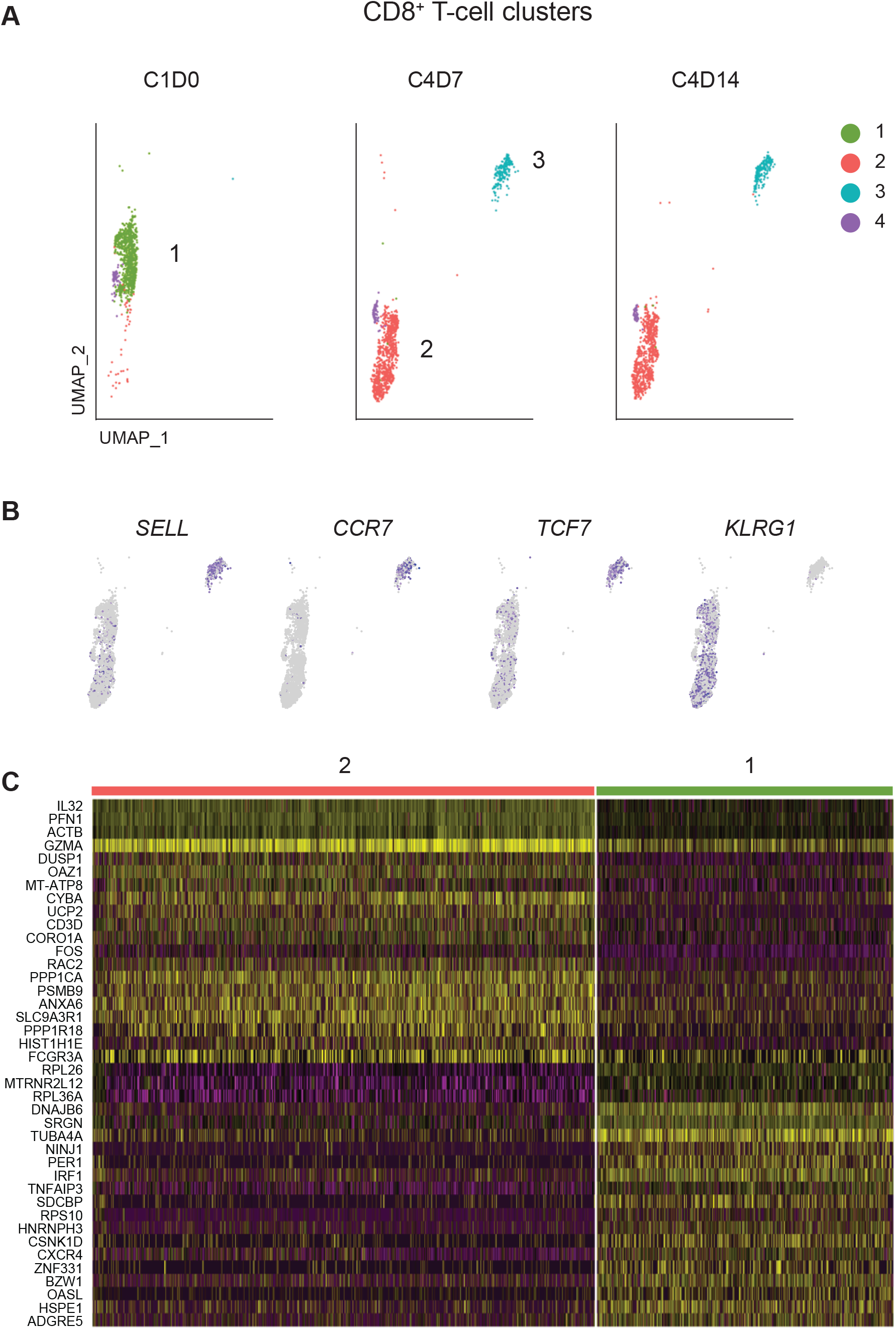
**A**, Uniform Manifold Approximation and Projection (UMAP) plots of four different CD8^+^ T-cell clusters identified in a responding CRC patient receiving FP-1305. Clusters 2 and 3 expanded during treatment cycle 4 (C4). **B**, Cluster #3 represents resting CD8^+^ T-cells, which can be either naïve or circulating central memory cells according to *SELL, CCR7* and *TCF7* gene expression. **C**, Gene expression differences between clusters 1 and 2.

**Supplementary Table 1.**
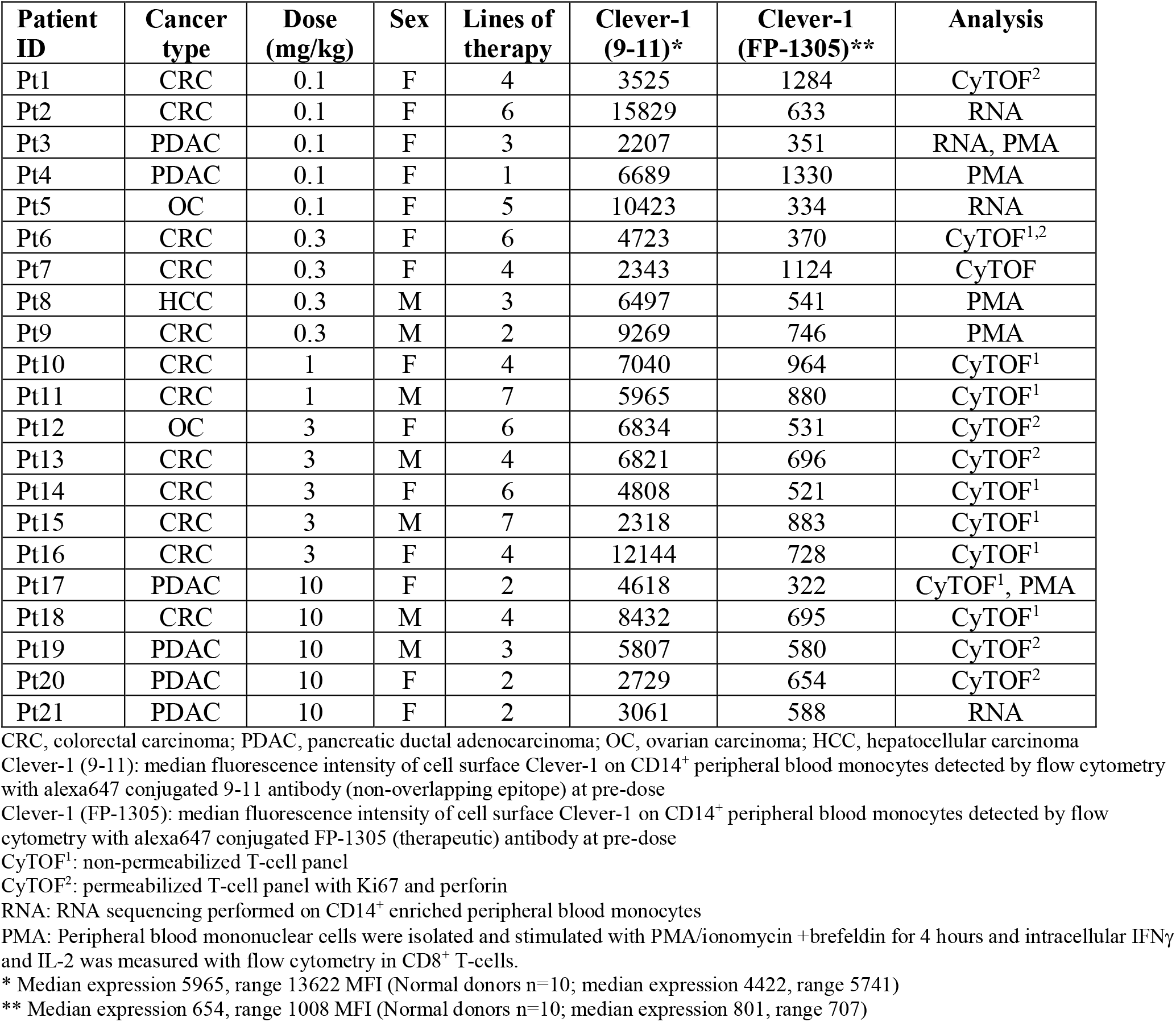
Characteristics of patient samples analyzed for systemic responses.

| Patient ID | Cancer type | Dose (mg/kg) | Sex | Lines of therapy | Clever-1 (9-11)* | Clever-1 (FP-1305)** | Analysis |
| --- | --- | --- | --- | --- | --- | --- | --- |
| Pt1 | CRC | 0.1 | F | 4 | 3525 | 1284 | CyTOF<sup>2</sup> |
| Pt2 | CRC | 0.1 | F | 6 | 15829 | 633 | RNA |
| Pt3 | PDAC | 0.1 | F | 3 | 2207 | 351 | RNA, PMA |
| Pt4 | PDAC | 0.1 | F | 1 | 6689 | 1330 | PMA |
| Pt5 | OC | 0.1 | F | 5 | 10423 | 334 | RNA |
| Pt6 | CRC | 0.3 | F | 6 | 4723 | 370 | CyTOF<sup>1,2</sup> |
| Pt7 | CRC | 0.3 | F | 4 | 2343 | 1124 | CyTOF |
| Pt8 | HCC | 0.3 | M | 3 | 6497 | 541 | PMA |
| Pt9 | CRC | 0.3 | M | 2 | 9269 | 746 | PMA |
| Pt10 | CRC | 1 | F | 4 | 7040 | 964 | CyTOF<sup>1</sup> |
| Pt11 | CRC | 1 | M | 7 | 5965 | 880 | CyTOF<sup>1</sup> |
| Pt12 | OC | 3 | F | 6 | 6834 | 531 | CyTOF<sup>2</sup> |
| Pt13 | CRC | 3 | M | 4 | 6821 | 696 | CyTOF<sup>2</sup> |
| Pt14 | CRC | 3 | F | 6 | 4808 | 521 | CyTOF<sup>1</sup> |
| Pt15 | CRC | 3 | M | 7 | 2318 | 883 | CyTOF<sup>1</sup> |
| Pt16 | CRC | 3 | F | 4 | 12144 | 728 | CyTOF<sup>1</sup> |
| Pt17 | PDAC | 10 | F | 2 | 4618 | 322 | CyTOF<sup>1</sup>, PMA |
| Pt18 | CRC | 10 | M | 4 | 8432 | 695 | CyTOF<sup>1</sup> |
| Pt19 | PDAC | 10 | M | 3 | 5807 | 580 | CyTOF<sup>2</sup> |
| Pt20 | PDAC | 10 | F | 2 | 2729 | 654 | CyTOF<sup>2</sup> |
| Pt21 | PDAC | 10 | F | 2 | 3061 | 588 | RNA |

CRC, colorectal carcinoma; PDAC, pancreatic ductal adenocarcinoma; OC, ovarian carcinoma; HCC, hepatocellular carcinoma
Clever-1 (9-11): median fluorescence intensity of cell surface Clever-1 on CD14<sup>+</sup> peripheral blood monocytes detected by flow cytometry with alexa647 conjugated 9-11 antibody (non-overlapping epitope) at pre-dose
Clever-1 (FP-1305): median fluorescence intensity of cell surface Clever-1 on CD14<sup>+</sup> peripheral blood monocytes detected by flow cytometry with alexa647 conjugated FP-1305 (therapeutic) antibody at pre-dose
CyTOF<sup>1</sup>: non-permeabilized T-cell panel
CyTOF<sup>2</sup>: permeabilized T-cell panel with Ki67 and perforin
RNA: RNA sequencing performed on CD14<sup>+</sup> enriched peripheral blood monocytes
PMA: Peripheral blood mononuclear cells were isolated and stimulated with PMA/ionomycin +brefeldin for 4 hours and intracellular IFNγ and IL-2 was measured with flow cytometry in CD8<sup>+</sup> T-cells.
\* Median expression 5965, range 13622 MFI (Normal donors n=10; median expression 4422, range 5741)
\*\* Median expression 654, range 1008 MFI (Normal donors n=10; median expression 801, range 707)

**Supplementary Table 2.** Cluster definition of PBMCs (pre-gated by viable singlets and CD45^+^CD3^+^)

| Cluster # | Name | Marker expression |
| --- | --- | --- |
| **1** | CD4 naive | CD4+ CD45RA- CCR7+ CD45RO- CD127- CD25- CCR6- CXCR3- |
| **2** | CD4 naive | CD4+ CD45RA+ CCR7+ CD45RO- CD127+ CD25- CCR6- CXCR3- |
| **3** | CD4 naive | CD4+ CD45RA- CCR7+ CD45RO- CD127+ CD25+ CCR6- CXCR3- |
| **4** | CD4 naive | CD4+ CD45RA- CCR7+ CD45RO- CD127+ CD25- CCR6+ CXCR3+ |
| **5** | CD4 naive | CD4+ CD45RA+ CCR7+ CD45RO- CD127+ CD25+ CCR6- CXCR3- |
| **6** | CD4 EFF | CD4+ CD45RA- CCR7- CD45RO- CD127- CD25- CCR6- CXCR3- |
| **7** | CD4 EM | CD4+ CD45RA- CCR7- CD45RO+ CD127+ CD25+ CCR6- CXCR3- |
| **8** | CD4 CM | CD4+ CD45RA- CCR7+ CD45RO+ CD127+ CD25+ CCR6+ CXCR3- |
| **9** | CD4 CM | CD4+ CD45RA- CCR7+ CD45RO+ CD127+ CD25+ CCR6- CXCR3- |
| **10** | CD4 Tregs | CD4+ CD45RA- CCR7+ CD45RO+ CD127- CD25+ CCR6- CXCR3- |
| **11** | CD8 naive | CD8+ CD45RA+ CCR7+ CD45RO- CD127+ CD25- CCR6- CXCR3- |
| **12** | CD8 naive | CD8+ CD45RA- CCR7+ CD45RO- CD127+ CD25- CCR6- CXCR3- |
| **13** | CD8 EFF | CD8+ CD45RA+ CCR7- CD45RO- CD127- CD25- CCR6- CXCR3- |
| **14** | CD8 EFF | CD8+ CD45RA- CCR7- CD45RO- CD127- CD25- CCR6+ CXCR3+ |
| **15** | CD8 EFF | CD8+ CD45RA+ CCR7- CD45RO- CD127- CD25- CCR6- CXCR3+ |
| **16** | CD8 EFF | CD8+ CD45RA+ CCR7- CD45RO- CD127- CD25- CCR6- CXCR3+ |
| **17** | CD8 EM | CD8+ CD45RA- CCR7- CD45RO+ CD127+ CD25- CCR6+ CXCR3- |
| **18** | CD8 EM | CD8+ CD45RA- CCR7- CD45RO+ CD127- CD25+ CXCR3+ CCR6- |
| **19** | CD4- CD8- | CD4- CD8- CD45RA+ CCR7+ CD45RO- CD127+ CD25- CCR6- CXCR3+ |
| **20** | CD4- CD8- | CD4- CD8- CD45RA+ CCR7- CD45RO- CD127- CD25- CCR6- CXCR3- |
| **21** | CD4- CD8- | CD4- CD8- CD45RA- CCR7- CD45RO- CD127+ CD25- CCR6- CXCR3+ |
| **22** | CD4+ CD8+ | CD4+ CD8+ CD45RA- CCR7+ CD45RO+ CD127+ CD25+ CCR6+ CXCR3+ |

**Supplementary Table 3.** Cluster definition of PBMCs (pre-gated by viable singlets and CD45^+^CD3^-^).

| Cluster # | Name | Marker expression |
| --- | --- | --- |
| 1 | Classical monocytes | CD16- CD14+ CD11b+ CD64+ CD11c+ HLA DR+ |
| 2 | Classical monocytes | CD16- CD14+ CD11b+ CD64+ CD11c+ HLA DR+ |
| 3 | Classical monocytes | CD16- CD14+ CD11b+ CD64+ CD11c+ HLA DR+ |
| 4 | Classical monocytes | CD16<sup>Int</sup> CD14+ CD11b+ CD64+ CD11c+ HLA DR+ |
| 5 | Classical monocytes | CD16- CD14+ CD11b+ CD64+ CD11c+ HLA DR+ |
| 6 | Classical monocytes | CD16- CD14+ CD11b+ CD64+ CD11c+ HLA DR- |
| 7 | Classical monocytes | CD16- CD14+ CD11b+ CD64+ CD11c+ HLA DR+ |
| 8 | Classical monocytes | CD16- CD14+ CD11b+ CD64+ CD11c+ HLA DR- |
| 9 | Classical monocytes | CD16- CD14+ CD11b+ CD64+ CD11c+ HLA DR- |
| 10 | Undifferentiated myeloid cells | CD16- CD14- CD11b+ CD64- CD11c- HLA DR- |
| 11 | Intermediate monocytes | CD16+ CD14+ CD11b+ CD64+ CD11c+ HLA DR- |
| 12 | Non classical monocytes | CD16+ CD14- CD11b+ CD64+ CD11c+ HLA DR+ |
| 13 | DCs | CD16- CD14- CD11b- CD64+ CD11c+ HLA DR++ |
| 14 | B cells | CD19+ HLA DR+ CD16- |
| 15 | B cells | CD19+ HLA DR+ CD16- |
| 16 | B cells | CD19+ HLA DR+ CD16- |
| 17 | B cells | CD19+ HLA DR+ CD16- |
| 18 | B cells | CD19+ HLA DR- CD16- |
| 19 | B cells | CD19+ HLA DR+ CD16- |
| 20 | B cells | CD19+ HLA DR+ CD16+ |
| 21 | NK cells | CD56+ CD16- |
| 22 | NK cells | CD56+ CD16- |
| 23 | NK cells | CD56+ CD16+ |
| 24 | NK cells | CD56+ CD16+ |
| 25 | Undefined | CD16- CD14- CD11b- CD64- CD11c- HLA DR+ |
| 26 | Undefined | Negative for all |

## References

Algars, A., Irjala, H., Vaittinen, S., Huhtinen, H., Sundström, J., Salmi, M., Ristamäki, R., and Jalkanen, S. (2012). Type and location of tumor-infiltrating macrophages and lymphatic vessels predict survival of colorectal cancer patients. Int J Cancer 131, 864–873.

Alloatti, A., Kotsias, F., Pauwels, A. M., Carpier, J. M., Jouve, M., Timmerman, E., Pace, L., Vargas, P., Maurin, M., Gehrmann, U., et al. (2015). Toll-like Receptor 4 Engagement on Dendritic Cells Restrains Phago-Lysosome Fusion and Promotes Cross-Presentation of Antigens. Immunity 43, 1087–1100.

Anders, S., Pyl, P. T., and Huber, W. (2015). HTSeq--a Python framework to work with high-throughput sequencing data. Bioinformatics 31, 166–169.

Björkström, N. K., Gonzalez, V. D., Malmberg, K. J., Falconer, K., Alaeus, A., Nowak, G., Jorns, C., Ericzon, B. G., Weiland, O., Sandberg, J. K., and Ljunggren, H. G. (2008). Elevated numbers of Fc gamma RIIIA+ (CD16+) effector CD8 T cells with NK cell-like function in chronic hepatitis C virus infection. J Immunol 181, 4219–4228.

Bono, P., Virtakoivu, R., Vaura, F., Jaakkola, P., Shetty, S., Thibault, A., De Jonge, M. J., Minchom, A., Ma, Y. T., Yap, C., et al. (2020). Immune activation in first-in-human anti-macrophage antibody (anti-Clever-1 mAb; FP-1305) phase I/II MATINS trial: Part 1 dose-escalation, safety and efficacy results. J Clin Oncol 38, (suppl; abstr 3097).

Cassetta, L., and Pollard, J. W. (2018). Targeting macrophages: therapeutic approaches in cancer. Nat Rev Drug Discov 17, 887–904.

Collins, M. P., and Forgac, M. (2020). Regulation and function of V-ATPases in physiology and disease. Biochim Biophys Acta Biomembr, 183341.

De Simone, G., Mazza, E. M. C., Cassotta, A., Davydov, A. N., Kuka, M., Zanon, V., De Paoli, F., Scamardella, E., Metsger, M., Roberto, A., et al. (2019). CXCR3 Identifies Human Naive CD8. J Immunol 203, 3179–3189.

Donadon, M., Torzilli, G., Cortese, N., Soldani, C., Di Tommaso, L., Franceschini, B., Carriero, R., Barbagallo, M., Rigamonti, A., Anselmo, A., et al. (2020). Macrophage morphology correlates with single-cell diversity and prognosis in colorectal liver metastasis. J Exp Med 217.

Doncheva, N. T., Morris, J. H., Gorodkin, J., and Jensen, L. J. (2019). Cytoscape StringApp: Network Analysis and Visualization of Proteomics Data. J Proteome Res 18, 623–632.

Eisenhauer, E. A., Therasse, P., Bogaerts, J., Schwartz, L. H., Sargent, D., Ford, R., Dancey, J., Arbuck, S., Gwyther, S., Mooney, M., et al. (2009). New response evaluation criteria in solid tumours: revised RECIST guideline (version 1.1). Eur J Cancer 45, 228–247.

Etzerodt, A., Rasmussen, M. R., Svendsen, P., Chalaris, A., Schwarz, J., Galea, I., Møller, H. J., and Moestrup, S. K. (2014). Structural basis for inflammation-driven shedding of CD163 ectodomain and tumor necrosis factor-α in macrophages. J Biol Chem 289, 778–788.

Gu, Z., Eils, R., and Schlesner, M. (2016). Complex heatmaps reveal patterns and correlations in multidimensional genomic data. Bioinformatics 32, 2847–2849.

Hollmén, M., Figueiredo, C. R., and Jalkanen, S. (2020). New tools to prevent cancer growth and spread: a ‘Clever’ approach. Br J Cancer.

Hudson, W. H., Gensheimer, J., Hashimoto, M., Wieland, A., Valanparambil, R. M., Li, P., Lin, J. X., Konieczny, B. T., Im, S. J., Freeman, G. J., et al. (2019). Proliferating Transitory T Cells with an Effector-like Transcriptional Signature Emerge from PD-1. Immunity 51, 1043-1058.e1044.

Irjala, H., Elima, K., Johansson, E. L., Merinen, M., Kontula, K., Alanen, K., Grenman, R., Salmi, M., and Jalkanen, S. (2003). The same endothelial receptor controls lymphocyte traffic both in vascular and lymphatic vessels. Eur J Immunol 33, 815–824.

Jahchan, N. S., Mujal, A. M., Pollack, J. L., Binnewies, M., Sriram, V., Reyno, L., and Krummel, M. F. (2019). Tuning the Tumor Myeloid Microenvironment to Fight Cancer. Front Immunol 10, 1611.

Karikoski, M., Marttila-Ichihara, F., Elima, K., Rantakari, P., Hollmén, M., Kelkka, T., Gerke, H., Huovinen, V., Irjala, H., Holmdahl, R., et al. (2014). Clever-1/stabilin-1 controls cancer growth and metastasis. Clin Cancer Res 20, 6452–6464.

Kidani, Y., and Bensinger, S. J. (2012). Liver X receptor and peroxisome proliferator-activated receptor as integrators of lipid homeostasis and immunity. Immunol Rev 249, 72–83.

Kim, D., Langmead, B., and Salzberg, S. L. (2015). HISAT: a fast spliced aligner with low memory requirements. Nat Methods 12, 357–360.

Kimball, A. K., Oko, L. M., Bullock, B. L., Nemenoff, R. A., van Dyk, L. F., and Clambey, E. T. (2018). A Beginner’s Guide to Analyzing and Visualizing Mass Cytometry Data. J Immunol 200, 3–22.

Kratchmarov, R., Magun, A. M., and Reiner, S. L. (2018). TCF1 expression marks self-renewing human CD8. Blood Adv 2, 1685–1690.

Krämer, A., Green, J., Pollard, J., and Tugendreich, S. (2014). Causal analysis approaches in Ingenuity Pathway Analysis. Bioinformatics 30, 523–530.

Kzhyshkowska, J., Gratchev, A., and Goerdt, S. (2006). Stabilin-1, a homeostatic scavenger receptor with multiple functions. J Cell Mol Med 10, 635–649.

Love, M. I., Huber, W., and Anders, S. (2014). Moderated estimation of fold change and dispersion for RNA-seq data with DESeq2. Genome Biol 15, 550.

Melero, I., Rouzaut, A., Motz, G. T., and Coukos, G. (2014). T-cell and NK-cell infiltration into solid tumors: a key limiting factor for efficacious cancer immunotherapy. Cancer Discov 4, 522–526.

Mellacheruvu, D., Wright, Z., Couzens, A. L., Lambert, J. P., St-Denis, N. A., Li, T., Miteva, Y. V., Hauri, S., Sardiu, M. E., Low, T. Y., et al. (2013). The CRAPome: a contaminant repository for affinity purification-mass spectrometry data. Nat Methods 10, 730–736.

Morris, J. H., Apeltsin, L., Newman, A. M., Baumbach, J., Wittkop, T., Su, G., Bader, G. D., and Ferrin, T. E. (2011). clusterMaker: a multi-algorithm clustering plugin for Cytoscape. BMC Bioinformatics 12, 436.

Mou, D., Espinosa, J., Lo, D. J., and Kirk, A. D. (2014). CD28 negative T cells: is their loss our gain? Am J Transplant 14, 2460–2466.

Nagy, L., Szanto, A., Szatmari, I., and Széles, L. (2012). Nuclear hormone receptors enable macrophages and dendritic cells to sense their lipid environment and shape their immune response. Physiol Rev 92, 739–789.

Palani, S., Elima, K., Ekholm, E., Jalkanen, S., and Salmi, M. (2016). Monocyte Stabilin-1 Suppresses the Activation of Th1 Lymphocytes. J Immunol 196, 115–123.

Palani, S., Maksimow, M., Miiluniemi, M., Auvinen, K., Jalkanen, S., and Salmi, M. (2011). Stabilin-1/CLEVER-1, a type 2 macrophage marker, is an adhesion and scavenging molecule on human placental macrophages. Eur J Immunol 41, 2052–2063.

Rantakari, P., Patten, D. A., Valtonen, J., Karikoski, M., Gerke, H., Dawes, H., Laurila, J., Ohlmeier, S., Elima, K., Hübscher, S. G., et al. (2016). Stabilin-1 expression defines a subset of macrophages that mediate tissue homeostasis and prevent fibrosis in chronic liver injury. Proc Natl Acad Sci U S A 113, 9298–9303.

Reddy, M. P., Kinney, C. A., Chaikin, M. A., Payne, A., Fishman-Lobell, J., Tsui, P., Dal Monte, P. R., Doyle, M. L., Brigham-Burke, M. R., Anderson, D., et al. (2000). Elimination of Fc receptor-dependent effector functions of a modified IgG4 monoclonal antibody to human CD4. J Immunol 164, 1925–1933.

Robinson, M. D., and Oshlack, A. (2010). A scaling normalization method for differential expression analysis of RNA-seq data. Genome Biol 11, R25.

Shannon, P., Markiel, A., Ozier, O., Baliga, N. S., Wang, J. T., Ramage, D., Amin, N., Schwikowski, B., and Ideker, T. (2003). Cytoscape: a software environment for integrated models of biomolecular interaction networks. Genome Res 13, 2498–2504.

Sharma, P., Hu-Lieskovan, S., Wargo, J. A., and Ribas, A. (2017). Primary, Adaptive, and Acquired Resistance to Cancer Immunotherapy. Cell 168, 707–723.

Szklarczyk, D., Morris, J. H., Cook, H., Kuhn, M., Wyder, S., Simonovic, M., Santos, A., Doncheva, N. T., Roth, A., Bork, P., et al. (2017). The STRING database in 2017: quality-controlled protein-protein association networks, made broadly accessible. Nucleic Acids Res 45, D362–D368.

Viitala, M. K., Virtakoivu, R., Tadayon, S., Rannikko, J., Jalkanen, S., and Hollmén, M. (2019). Immunotherapeutic Blockade of Macrophage Clever-1 Reactivates the CD8+ T Cell Response Against Immunosuppressive Tumors. Clin Cancer Res.

Wang, L., Feng, Z., Wang, X., and Zhang, X. (2010). DEGseq: an R package for identifying differentially expressed genes from RNA-seq data. Bioinformatics 26, 136–138.

Wu, T. D., Madireddi, S., de Almeida, P. E., Banchereau, R., Chen, Y. J., Chitre, A. S., Chiang, E. Y., Iftikhar, H., O’Gorman, W. E., Au-Yeung, A., et al. (2020). Peripheral T cell expansion predicts tumour infiltration and clinical response. Nature 579, 274–278.

Zhang, Y., Reynolds, J. M., Chang, S. H., Martin-Orozco, N., Chung, Y., Nurieva, R. I., and Dong, C. (2009). MKP-1 is necessary for T cell activation and function. J Biol Chem 284, 30815–30824.

